# AI Chatbots Versus Human Healthcare Professionals: A Systematic Review and Meta-Analysis of Empathy in Patient Care

**DOI:** 10.1101/2025.06.09.25329258

**Authors:** Alastair Howcroft, Amber Bennett-Weston, Ahmad Khan, Joseff Griffiths, Simon Gay, Jeremy Howick

**Author notes:** Corresponding Author Alastair Howcroft, Address for correspondence: Stoneygate Centre for Empathic Healthcare, Leicester Medical School, University of Leicester, George Davies Centre, Lancaster Rd, Leicester LE1 7HA, United Kingdom.

## Abstract

**Background:** Empathy is widely recognised for improving patient outcomes ranging from reduced pain and anxiety to improved patient satisfaction, and its absence can cause harm. Meanwhile, use of artificial intelligence (AI)–based chatbots in healthcare is rapidly expanding, with one in five general practitioners (GPs) using generative AI to assist with tasks such as writing letters. Several studies suggest that these AI-based technologies are sometimes more empathic than human healthcare professionals (HCPs). However, the evidence in this area is mixed and has not been synthesised.

**Objective:** To conduct a systematic review of studies that compare empathy of AI technologies with human HCP empathy.

**Methods:** We searched multiple databases for studies comparing AI chatbots using large language models (e.g., GPT-3.5, GPT-4) with human HCPs on empathy measures. We assessed risk of bias with ROBINS-I and synthesised findings using random-effects meta-analysis where feasible, whilst avoiding double counting.

**Results:** Our search identified 15 studies (2023–2024). Thirteen studies reported statistically significantly higher empathy ratings for AI, with only two studies situated in dermatology favouring human responses. Meta-analysis of 13 studies with data suitable for pooling, all utilising ChatGPT-3.5/4, showed a standardised mean difference (SMD) of 0.87 (95% CI, 0.54–1.20) favouring AI (p<0.00001).

**Conclusion:** Our findings indicate that, in text-only scenarios, AI chatbots are frequently perceived as more empathic than human HCPs – equivalent to an increase of approximately two points on a 10-point empathy scale. Future research should validate these findings with direct patient evaluations and assess whether emerging voice-enabled AI systems can deliver similar empathic advantages.

## 1. Introduction

Empathic healthcare is well recognised for its positive impact on patient quality of life, satisfaction with care, and reduction of pain and psychological distress [1]. With recent advances in artificial intelligence (AI) technologies, chatbots are increasingly being integrated into patient care, even replacing human practitioner roles at times. For example, Wysa – a digital therapist – has been used by over 117,000 patients across 31 NHS Talking Therapy services, according to Wysa’s official website [2]. These AI systems can interact with patients through text or speech, providing information, monitoring symptoms, offering support, and fulfilling other roles historically provided by human healthcare professionals (HCPs) [3]. It has also been reported that 20% of UK general practitioners (GPs) now use generative AI like ChatGPT for tasks such as assistance with writing patient correspondence [4]. However, despite the growing use of these technologies [5], there are concerns about whether AI chatbots can be as empathic as human HCPs [6], and thus leverage the benefits of empathic care for patients [7]. Such doubts align with the 2019 Topol Review, a UK government-commissioned roadmap for NHS technology, which concluded that ‘empathy and compassion’ remain ‘essential human skill[s] that AI cannot replicate’ [8].

Whilst individual studies have explored the potential of AI technologies to display empathic behaviours [9], the results are heterogeneous. Some studies suggest that AI bots can produce responses with higher levels of perceived empathy [10], whilst others cite issues suggesting the contrary, maintaining that they lack the warmth, nuanced understanding, and ability to form deep emotional bonds inherent in human interactions [11]. Existing literature reviews on the use of AI bots have primarily focused on disease diagnosis, clinical efficiency, ethical considerations, and their integration into healthcare systems [5, 12–14] and factual accuracy of medical advice compared to humans [15].

The lack of a synthesis of studies comparing AI technology with human empathy represents a significant gap in the literature. Such a synthesis can pave the way for more informed decisions about the integration of AI technologies into patient care, ensuring that such technological advancements contribute to (and don’t detract from) the positive patient outcomes associated with traditional empathic healthcare.

## 2. Methods

The review is reported according to the PRISMA 2020 Checklist.

### 2.1 Eligibility criteria

Studies were eligible if they empirically compared empathy between AI chatbots and human HCPs. Eligible study designs included quantitative or qualitative studies involving real patients, healthcare users, or authentic patient-generated data, such as emails, portal messages, or public forum posts. We included participants of any age, gender, or demographic engaging in formal or informal healthcare interactions. AI interventions were restricted to conversational agents using Large Language Models (LLMs), such as GPT-3/4, Claude, or Gemini, capable of unscripted dialogue. Studies comparing AI empathy directly to human HCP empathy were required.

Exclusions included simulated or hypothetical patient scenarios without authentic patient-generated data, healthy volunteers without healthcare needs, and interactions outside healthcare contexts (e.g., education, customer service). We also excluded studies utilising rule-based or scripted AI systems, opinion pieces, theoretical discussions, editorials, commentaries, and reviews lacking original empirical data.

### 2.2 Information sources

We searched PubMed, Cochrane Library, Embase, PsycINFO, CINAHL, Scopus, and IEEE Xplore from inception to 11 November 2024. ClinicalTrials.gov, ICTRP, and ISRCTN were searched for completed studies with published results. Reference lists of included studies and relevant reviews were screened, and grey literature was searched via Google Scholar on 12 November 2024.

### 2.3 Search strategy

The search strategy focused on three key groups of terms: empathy, AI technologies, and HCPs. Relevant keywords and index terms were identified through preliminary searches in IEEE Xplore and PubMed, supported by consultation with an academic librarian. Broad terms for AI and HCPs captured diverse technologies and roles, whereas empathy terms (“empathy”, “empathic”, “empathetic”, “compassion”, and derivatives) were narrower to maintain specificity. Groups were combined with “OR” within categories and “AND” across categories. The full strategy is detailed in Appendix A.

### 2.4 Selection process

The primary reviewer independently screened all titles and abstracts to ascertain which studies to include or exclude based on the predefined eligibility criteria. The abstracts were divided between two secondary reviewers, who independently screened their assigned portions. Discrepancies between the primary reviewer and the specific secondary reviewer who assessed the abstract were resolved through discussion. Similarly, this process was repeated for the full-text screening stage.

### 2.5 Data collection process

All identified records were imported into EndNote [16] for organisation and removal of duplicates. Study selection was managed using Rayyan [17]. The primary reviewer developed and independently completed the data extraction form, with two secondary reviewers independently extracting data on assigned portions, guided initially by an example form to ensure consistency.

### 2.6 Data Items

We collected data on study design, participants, settings, AI interventions, human comparators, empathy measures, and key findings related to empathy.

### 2.7 Synthesis Methods

Despite heterogeneity, particularly in terms of empathy measurement instruments (ranging from custom Likert-type scales to qualitative coding) and empathy evaluators (patient proxies, medical students, etc.), studies were sufficiently similar for meta-analysis due to shared structural elements. All studies directly compared AI-generated responses to human HCPs using blinded evaluations (except one not stating blinding), and all but that same study quantitatively provided empathy results (mostly via single-item Likert scales), enabling relative effect size calculations. All comparisons involved language-based interactions, primarily text (one study used audio-converted text). We also narratively synthesised the results (Appendix C). We organised studies by the large language model (GPT-3.5, GPT-4, other models) and then compared outcomes against its HCP comparators (e.g., physicians, nurses). Where available, we reported empathy scores, mean differences, p-values (for statistical significance), and other relevant summary statistics.

### 2.8 Reporting bias assessment

Two reviewers independently assessed the risk of bias. The ROBINS-I tool [18] was used to assess the risk of bias (RoB) across the 15 studies, given that most were non-randomised designs. Discrepancies were resolved through discussion, and with a third reviewer where necessary.

### 2.9 Meta-analysis

We conducted a single meta-analysis with two subgroups (GPT-3.5 and GPT-4) due to their prevalence in the literature (five and ten appearances, respectively; other models appeared once). To prevent double-counting, we excluded additional AI arms from multi-model studies and entirely omitted studies evaluating both GPT versions on the same dataset. We extracted SMDs with 95% confidence intervals (CIs) and constructed random-effects models to pool effect sizes. Subgroup differences were examined by comparing pooled effect estimates, with p<0.05 considered statistically significant. Analyses and plots used Review Manager (RevMan) 5.4 [19].

## 3. Results

### 3.1 Study Selection

The search identified 987 unique results that were screened by title/abstract and by full text. Following title and abstract screening, 34 articles were included. During full-text screening, an additional 19 studies were excluded (Appendix B). Ultimately, 15 studies met the inclusion criteria for this systematic review (see Figure 1).

**Fig. 1.**
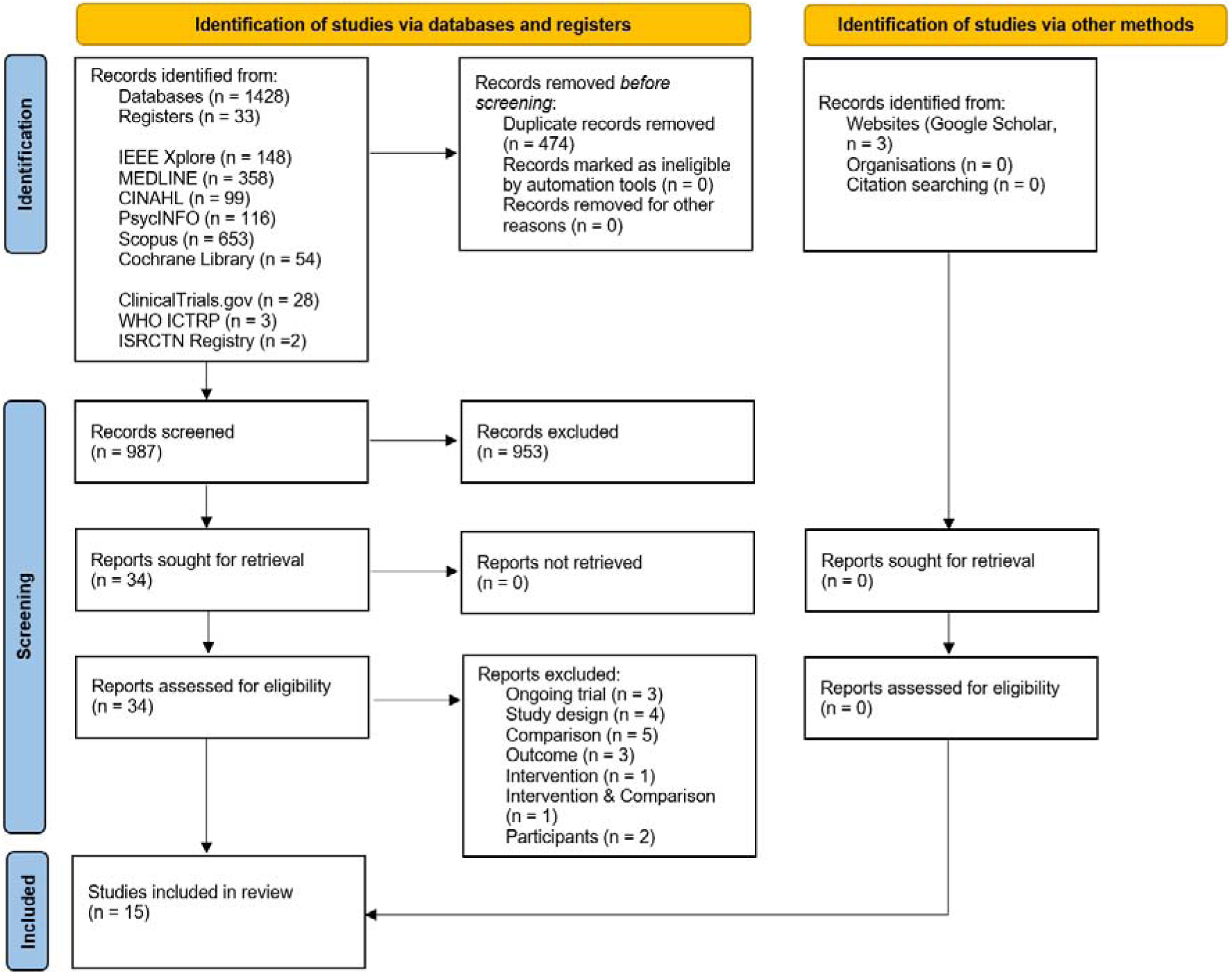
PRISMA Flow diagram.

### 3.2 Risk of bias

Overall, nine studies had a moderate risk of bias, and six had a serious risk of bias (Appendix D). Seven used curated patient queries, potentially introducing selection bias. Four relied on Reddit communities [10, 20–22] – an informal forum often serving socioeconomically disadvantaged users [23] – leading to serious concerns about representativeness, thoroughness, and confounding. Other serious cases employed supervised designs (i.e., “Wizard-of-Oz”, AI–nurse collaboration) [24, 25], complicating isolation of chatbot performance. Heterogeneity arose from diverse empathy raters (e.g., patient proxies, clinicians, psychology trainees), complicating comparisons because perceptions of empathy may vary by evaluator background. Finally, 14 of 15 studies assessed empathy using non-validated methods (e.g., single-item custom Likert scales), rather than a validated instrument (e.g., CARE) [26], limiting standardisation and heightening subjectivity.

### 3.3 Characteristics of Included Studies

Fourteen of the fifteen studies included in the review were published in 2024; the remaining study was published in 2023 [10].

#### 3.3.1 Types of health concerns and speciality

The studies covered diverse health conditions and specialities (see Table 1). Four involved non-specific medical concerns, reflecting routine and general patient inquiries. These included outpatient queries across multiple departments [25], general health questions from social media [10, 27], and internal medicine patient portal interactions, including lab results and administrative requests [28]. Other studies addressed specialised clinical conditions, such as dermatology [24, 29], oncology [20], and thyroid conditions [30]. Chronic diseases were also covered, including systemic lupus erythematosus [31] and multiple sclerosis [26], alongside reconstructive surgery [32] and complete blood count lab result interpretation [21]. Two addressed mental and neurodevelopmental health, where Yonatan-Leus and Brukner [22] examined mental health support inquiries, and He, Zhang, Jin, Zhou, Zhang and Xia [33] focused on autism-related queries. Finally, one service-oriented study examined patient complaints from various departments [34].

**Table 1:**
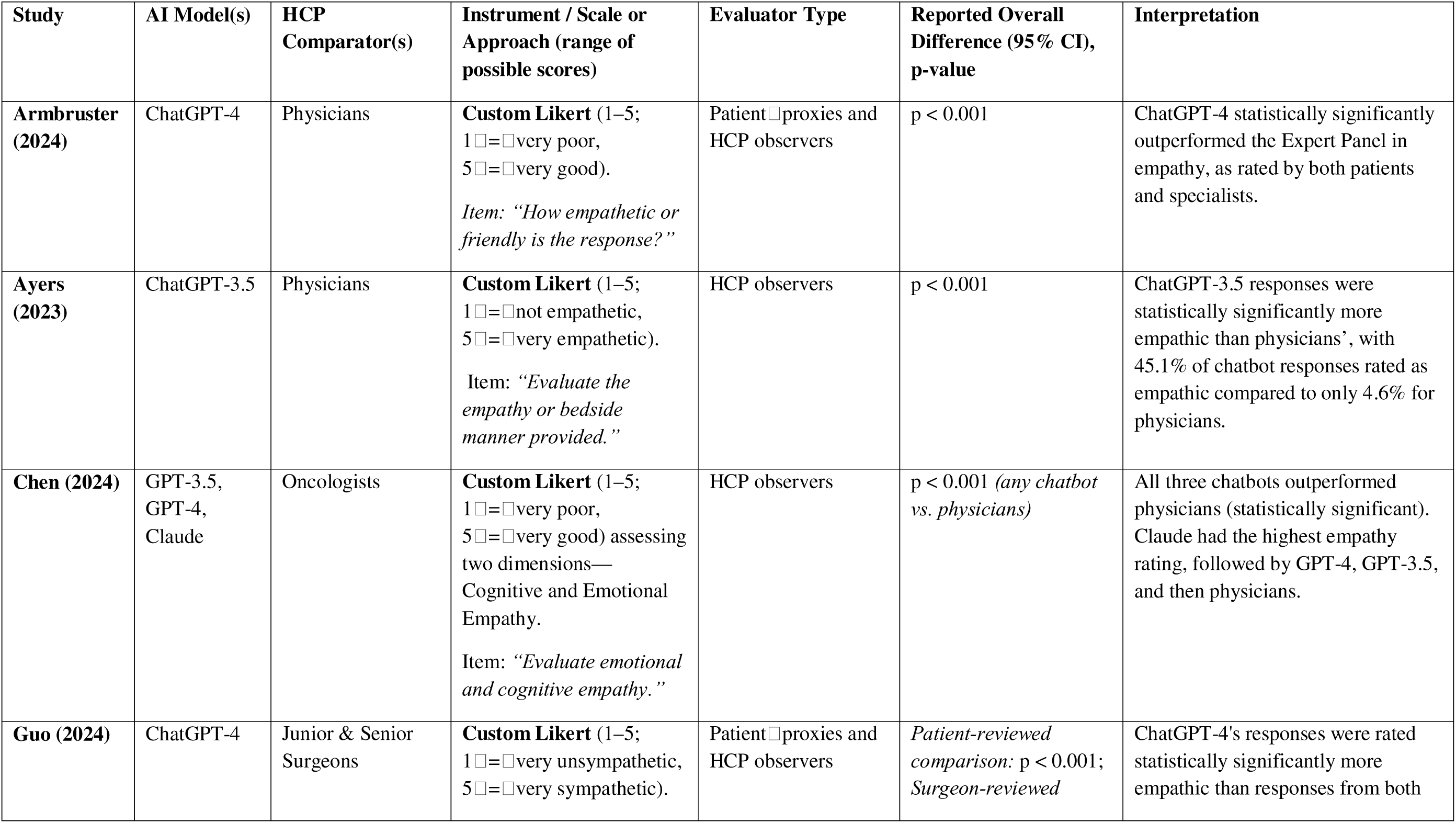
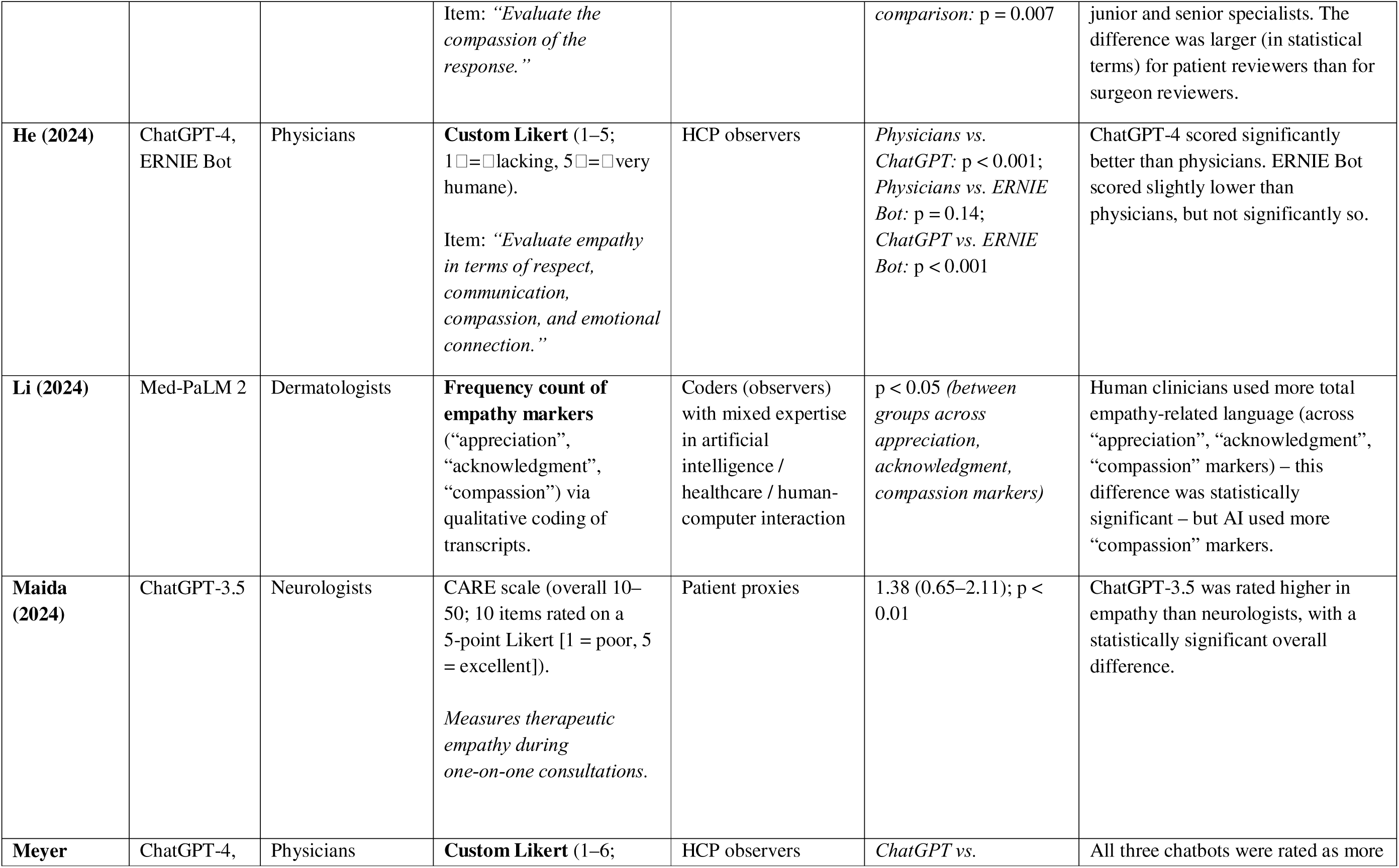
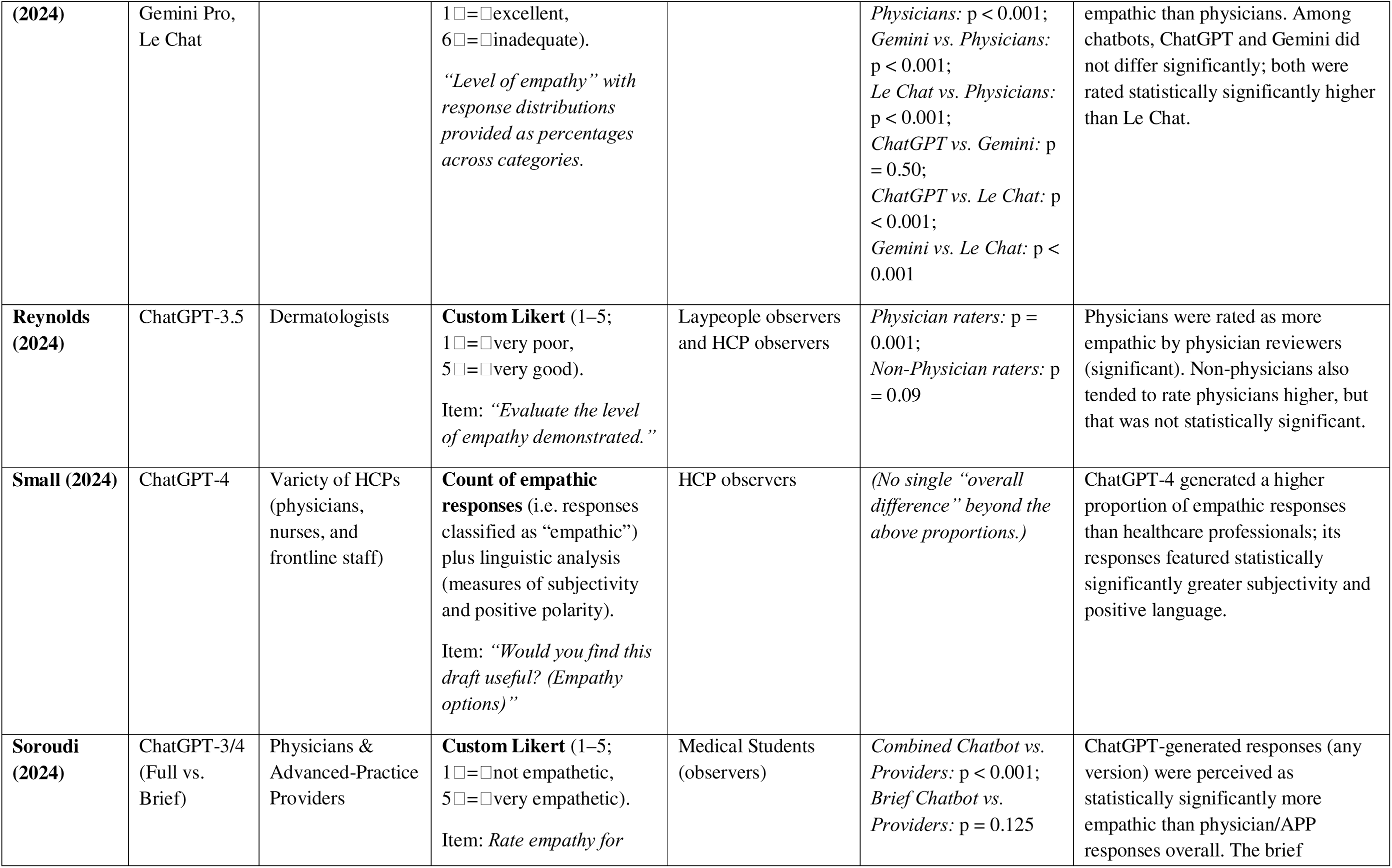
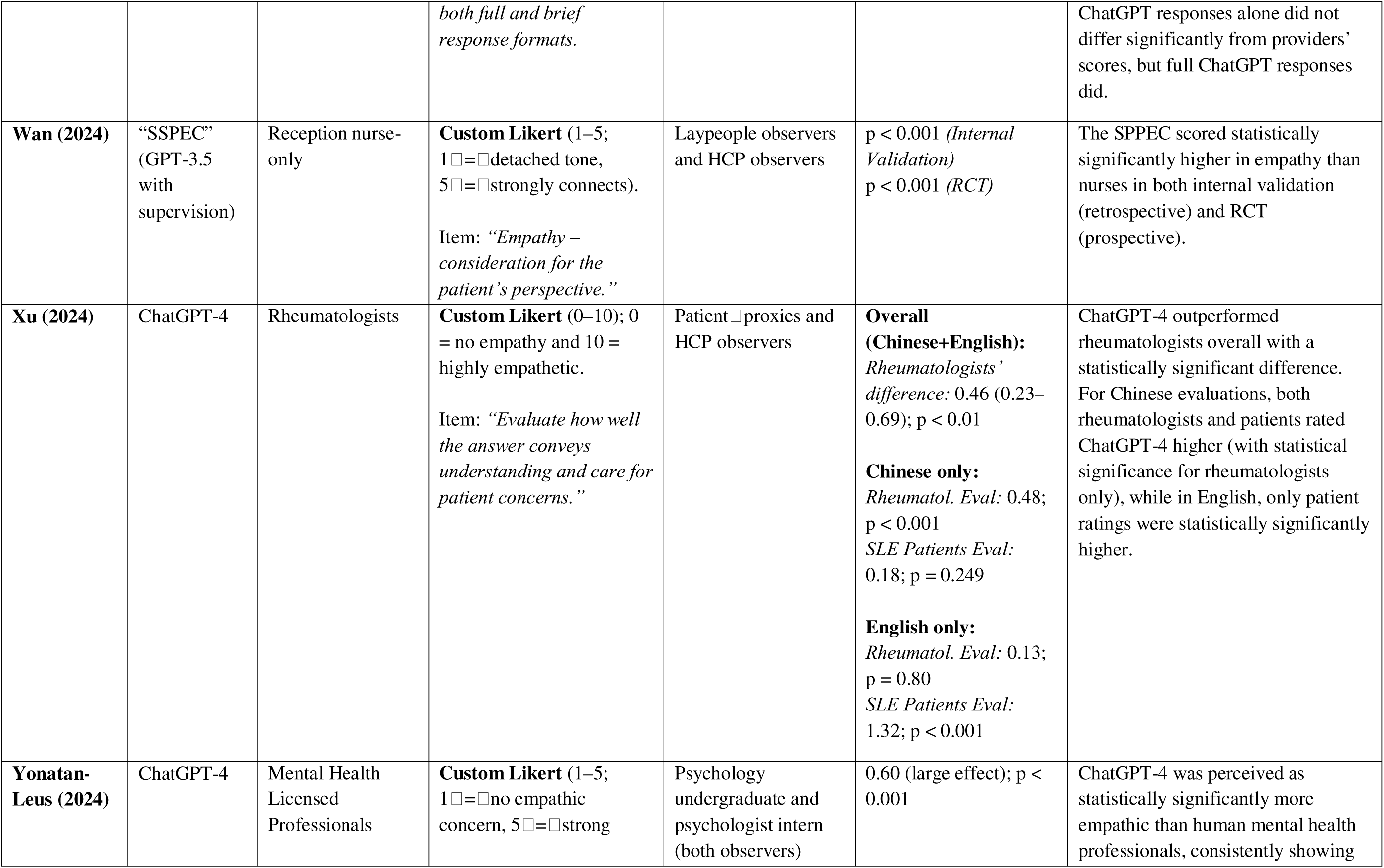
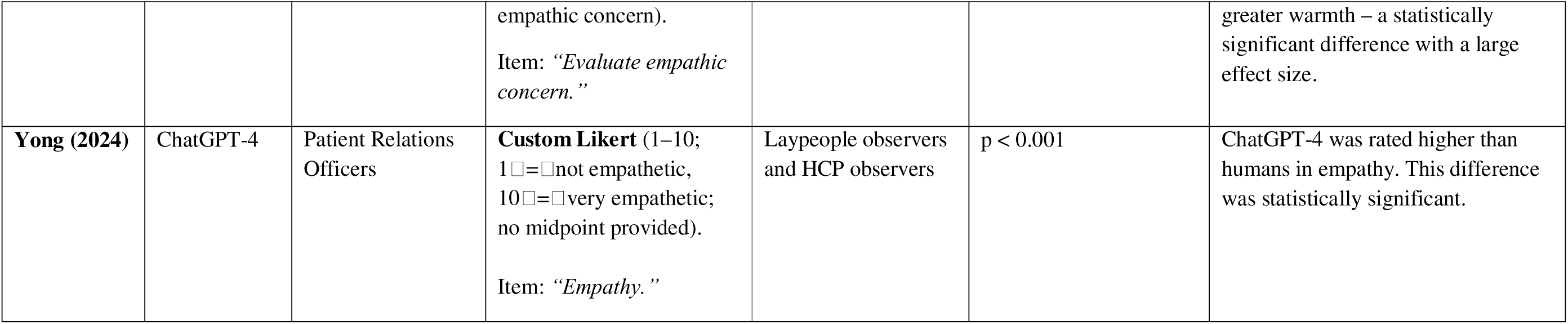
Empathy Comparisons Between AI Chatbots and Human Healthcare Practitioners – Summary of Findings Across Studies.

#### 3.3.2 Human HCP Comparators

The included studies compared AI-generated responses to a variety of human comparators. Two studies compared AI responses to those from surgeons [30, 32], while six studies involved comparisons with physicians specialising in specific fields [10, 20, 24, 26, 29, 33]. Three studies compared AI responses to physicians with no specified specialties [10, 21, 27]. Other comparisons included advanced practice providers in one study [32], nurses and frontline staff in one study [28], and reception nurses in another [25]. Mental health licensed professionals were the comparators in one study [22], whilst patient relations officers were included in another [34].

#### 3.3.3 Interaction Modalities and AI Models

All but one of the included studies relied exclusively on text-based interaction with the AI system. In one study, patient speech was transcribed (through a real-time voice transcribing software) into text for the LLM (GPT-3.5) and then the AI’s text response was converted back into audio [25]. One study allowed patients to upload images, however they were described textually by a human intermediary before input [24]. All but one study [24] used versions of GPT (general-purpose language models). Three studies also evaluated GPT alongside other LLMs: one with ERNIE Bot [33], one with Claude [20], and one with Gemini Pro and Le Chat [21]. The exception was a study exclusively using Med-PaLM2, a model specifically designed for medical question-answering tasks [24].

#### 3.3.4 Empathy Measurement Tools

All, bar one study, relied on unvalidated or custom tools for measuring empathy; the exception used the Consultation and Relational Empathy (CARE) scale [26], which is a validated instrument. Eight studies relied on single-item 1–5 Likert scales, where responses were rated from “not empathetic” to “very empathetic” or similar descriptors [10, 22, 25, 27, 29, 30, 32, 33]. One additional study used a 1–5 Likert scale, however, assessed separate scores for cognitive and emotional empathy [20]. Two studies used single-item 0/1–10 scales [31, 34] and another employed a single-item 1–6 scale [21]. Another study used a thematic coding framework, identifying and counting empathy-related statements – appreciation, acknowledgment, and compassion – as part of content analysis [24]. Additionally, one study measured empathy conditionally: reviewers first determined if responses (AI or human generated) were usable, then selected empathy as a reason for usefulness if applicable, rather than using any scale [28].

#### 3.3.5 Query Sources

The studies took patient inquiries, which were responded to by humans, AI models, or both, from a variety of sources. Seven studies utilised historical emails or message logs from private medical records [13, 26, 28–32, 34], six relied on publicly available questions from platforms like Reddit and other online forums [10, 20–22, 27, 33], one collected real-time chat transcripts (where messages are exchanged instantly) [24], and one involved in-person reception interactions in a hospital setting [25].

#### 3.3.6 How empathy was assessed

Across these studies, *all* evaluators functioned as “observers” (not those asking/answering the inquiries) but varied in their backgrounds. One study used patient proxies [26]; five employed HCPs [10, 20, 25, 28, 30]; three combined patient proxies and HCPs [27, 30, 31]; three involved laypeople alongside HCPs [25, 29, 34]; one used medical students [32]; and one featured a psychology undergraduate and psychologist intern [22]. All reviewers in these studies were blinded to whether the responses were generated by AI or humans. The additional study relied on researchers or coders with mixed expertise in healthcare, conversational AI, and human-computer interaction, and blinding was not mentioned [24].

## 4. Results of Included Studies

Table 1 summarises the characteristics of the 15 included studies, with statistical results reported as presented in the studies (sub-scores, overall means, etc.). Detailed study-specific results, including granular metrics and model comparisons, are provided in Appendix E.

## 5. Results of Syntheses

### 5.1 Overall Summary of Included Comparisons

In 13 out of the 15 comparisons, one or more AI chatbots demonstrated a statistically significant advantage in projection of empathy over human healthcare practitioners [10, 20–22, 25–28, 30–34] (see Figure 2). In one of these 13 comparisons, one AI model (ERNIE Bot) of the two assessed did not show a statistically significant difference from humans, but the other model (GPT-4) within the same study did outperform ERNIE Bot, demonstrating a statistically significant result [33]. In the remaining two studies, both involving dermatology, human dermatologists outperformed AI (Med-PaLM 2 and ChatGPT-3.5) in perceived empathy [24, 29].

**Fig. 2.**
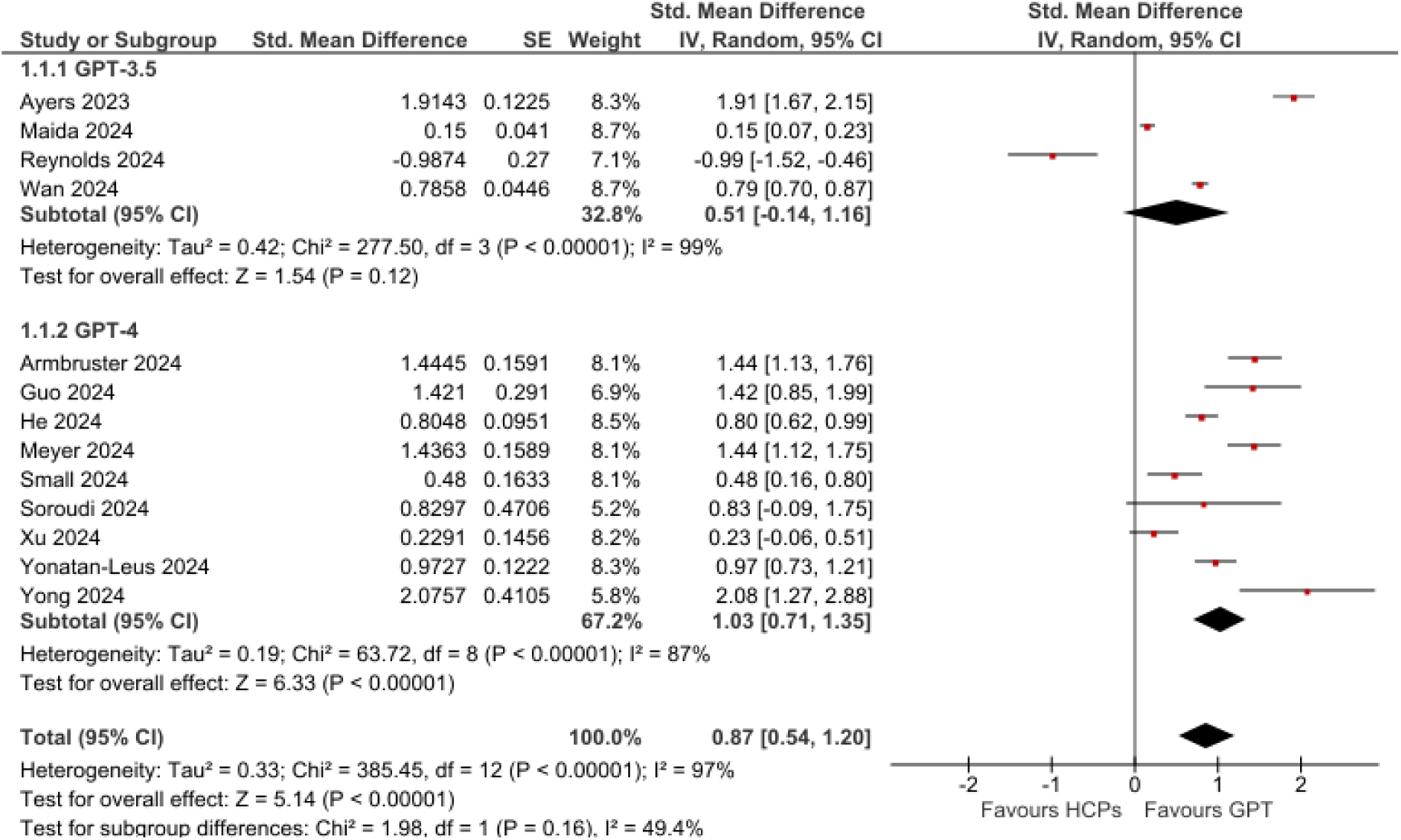
Forest plot comparing empathy ratings: GPT-3.5 vs. human practitioners and GPT-4 vs. human practitioners (excluding overlap).

We first present the results of our meta-analyses for GPT-3.5 and GPT-4. Afterwards, we briefly discuss the two studies that could not be included in the meta-analysis (n=1 to prevent double-counting [29], n=1 due to missing outcome data [24]) and other LLM arms (GPT-3, Gemini, Le Chat, and ERNIE Bot) beyond GPT-3.5/4. These additional models were excluded from the meta-analysis to avoid overlapping data. A full narrative synthesis of all models and studies are included in Appendix C.

### 5.2 Meta-Analyses of GPT-3.5 and GPT-4 Chatbots

Thirteen studies provided data suitable for meta-analysis. Overall, ChatGPT demonstrated significantly higher empathy than human practitioners (SMD 0.87, 95% CI 0.54–1.20; p<0.00001). Heterogeneity was moderate between GPT-3.5 and GPT-4 subgroups (I²=49.4%) but high across all studies (I²=97%). In Meyer et al. [21], the same 100 questions were tested with ChatGPT, Gemini, and Le Chat, risking double counting if pooled. Thus, it could not be pooled. As highlighted by Husssein, et al. [35], analyses of health record data are particularly prone to such double-counting errors. Similarly, Soroudi, et al. [32] compared GPT-3, GPT-4, and human HCPs; we retained only GPT-4 data to prevent double-counting. He, et al. [33] also tested ERNIE Bot and GPT-4 on the same dataset, so we included only GPT-4. Finally, Chen, et al. [20] simultaneously assessed GPT-3.5 and GPT-4 (and Claude) on the same dataset, creating an irresolvable conflict between our GPT-3.5 and GPT-4 subgroups; therefore, that study was omitted entirely from the pooled analysis. Li, et al. [24] evaluated Med-PaLM 2, but didn’t provide specific metrics, so was similarly excluded from the following meta-analysis.

### 5.3 Subgroup: GPT-3.5

Four studies provided data compatible with a GPT-3.5 subgroup meta-analysis (excluding Chen et al.). They reported empathy outcomes on validated (CARE) or custom Likert scales allowing calculation of standardized mean differences (SMDs). Pooled analysis (random-effects model) yielded an SMD of 0.51 (95% CI -0.14–1.16) favouring GPT-3.5 over human healthcare practitioners (I² = 99%), indicating high heterogeneity. This result was not statistically significant, however (p = 0.12), due to the one instance where HCPs scored higher in Reynolds, et al. [29].

Across four studies, GPT-3.5 outperformed human clinicians in three settings. Maida, et al. [26] reported an SMD of 0.15 (95% CI: 0.07–0.23) for neurologic inquiries, Wan, et al. [25] found an SMD of 0.79 (95% CI: 0.70–0.87) in an outpatient reception setting, and Ayers, et al. [10] observed an SMD of 1.91 (95% CI: 1.67–2.15) based on social media queries. In contrast, Reynolds, et al. [29] showed that for dermatology queries, human responses were rated higher (SMD −0.99, 95% CI: −1.52 to −0.46). We narratively synthesise these results in Appendix C.

### 5.4 Subgroup: GPT-4

Nine studies contributed data for the GPT-4 subgroup, yielding a pooled SMD of 1.03 (*95% CI* 0.71–1.35) in favour of GPT-4 (I² = 87%, Tau² = 0.19), again reflecting high heterogeneity. This result was statistically significant (p < 0.00001).

Across nine studies, GPT-4 consistently outperformed human clinicians. Guo, et al. [30] reported an SMD of 1.42 (95% CI: 0.85–1.99) for thyroid-related inquiries; Yonatan-Leus and Brukner [22] found an SMD of 0.97 (95% CI: 0.73–1.21) for mental health queries on social media; and Soroudi, et al. [32] observed an SMD of 0.83 (95% CI: −0.09–1.75) for breast reconstruction questions. Armbruster, et al. [27] showed an SMD of 1.44 (95% CI: 1.13–1.76) in a web-based setting, while He, et al. [33] reported an SMD of 0.80 (95% CI: 0.62–0.99) for autism-related inquiries. Xu, et al. [31] found an SMD of 0.23 (95% CI: −0.06–0.51) for systemic lupus erythematosus questions, and Yong, et al. [34] reported an SMD of 2.08 (95% CI: 1.27–2.88) for patient complaints. Additionally, Small, et al. [28] observed an SMD of 0.48 (95% CI: 0.16–0.80) for outpatient internal medicine queries, and Meyer, et al. [21] reported an SMD of 1.44 (95% CI: 1.12–1.75) for laboratory-interpretation queries. We narratively synthesise these results in Appendix C.

### 5.5 Subgroup Comparison: GPT-3.5 vs. GPT-4

When examining GPT-3.5 and GPT-4 side by side, GPT-4 consistently outperformed human clinicians in empathy, whilst GPT-3.5 showed mixed results and failed to demonstrate a statistically significant advantage. A subgroup analysis testing for differences between GPT-3.5 and GPT-4 revealed no statistically significant difference (p = 0.16). Thus, while GPT-4 achieved more consistent results, the current data do not conclusively indicate that it is definitively more empathic than GPT-3.5 (Figure 2). The pooled SMD of both models (n = 13 pooled studies) was 0.87 (95% CI: 0.54–1.20), with both GPT-3.5 and GPT-4 collectively demonstrating statistically significantly (p < 0.00001) higher empathy ratings than HCPs.

### 5.6 Non-Meta-Analysed Results

Across the non-metalJanalysed studies, Chen, et al. [20] found that GPT-3.5, GPT-4, and Claude all outperformed oncologists on oncology queries – with GPT-4 scoring highest. Li, et al. [24] showed that, while human clinicians used more empathy-related language overall, Med-PaLM 2 demonstrated a slight advantage in expressing compassion. Meyer, et al. [21] reported that both Gemini Pro and Le Chat (Mistral Large) were rated significantly higher than physicians, whereas He, et al. [33] found that ERNIE Bot slightly underperformed compared to clinicians. Finally, Soroudi, et al. [32] observed that GPT-3’s “full” format outperformed its “brief” format, with both formats exceeding providers’ ratings. For full details, please refer to Appendix C.

## 6. Discussion

### 6.1 Interpretation of Results

This is the first systematic review, we are aware of, that compares the empathy of AI chatbots with human practitioners. Our meta-analysis of 13 studies shows that ChatGPT has a 73% likelihood of being perceived as more empathic than a human practitioner in a head-to-head matchup, using text-based interactions (representing the probability of superiority). This finding was across multiple clinical specialties and various evaluative methods. This would be roughly equivalent to a difference of 2 points on a 10-point scale. An important implication is that text-based AI-driven interactions are unlikely to cause harm through deficits in empathy, aligning with broader evidence supporting AI’s potential to enhance healthcare engagement, quality, and efficiency [3].

### 6.2 Limitations

This study has several limitations. Firstly, all but one study (which converted AI-generated text to audio) [25], analysed text-based interactions. This is a problem because empathy in healthcare consultations often relies on both verbal and non-verbal cues (e.g., nodding and learning forward) [36], and because text-based communication represents a relatively small portion of healthcare interactions [37]. That being said, healthcare practitioners are increasingly using purely text-based communication, suggesting that, at least in these cases, our results are relevant to actual practice [38]. Another limitation is that the included studies evaluated empathy from proxy measures rather than the perspectives of the patients directly receiving care. Given that HCP and direct care recipients’ empathy ratings have been shown to differ [39, 40], it is possible that patient ratings would have been different.

Additionally, the potential gains in empathic communication must be weighed against ongoing concerns regarding the reliability of AI-driven clinical content [41]; any benefits in empathic delivery risk being overshadowed if the medical advice offered is inaccurate. Although empathy in healthcare (also called therapeutic or clinical empathy) is widely recognised as the ability to understand a patient’s emotions [42], it is important to note that the various studies define and operationalised empathy in their own ways.

Moreover, six of the included studies sourced patient questions and/or clinician responses from public forums. It could be that the AI chatbots (trained on vast internet datasets) [43] encountered the material during training, giving them an unfair advantage through test-set contamination. However, given the enormous volume of training data, any influence from specific queries is likely minimal. Also, models generate original, context-specific responses rather than reproducing exact copies [43]. However, this limitation could also work against the success of the AI chatbots: since human responses were generally less empathic, mimicking them would presumably reduce the chatbots’ relative empathy scores. Most (14/15) of the included studies focused on GPT-3/3.5 or GPT-4, which may limit generalisability to clinical practice. Widely deployed clinical chatbots (e.g., Wysa) often rely on proprietary models with distinct architectures and training data that could influence empathic communication. This variability raises questions about how well the findings translate to tools used in real-world care. Finally, all included studies were from 2024/2023; newer models such as GPT-4.5, released in February 2025 and claimed by OpenAI to demonstrate greater emotional intelligence [44], have not yet been evaluated in the literature. Another limitation is that the included studies exhibited substantial heterogeneity in outcome measures (with the included studies using a mix of singlelJitem Likert ratings, proportionlJbased assessments, validated and unvalidated tools), comparators (types of healthcare professional), and evaluators of empathy (including patient proxies, lay people, students, and HCPs, often in mixed combinations), complicating pooled inferences.

Relatedly, many studies lacked the detailed statistics (e.g., means, standard deviations, or standard errors) typically required for SMD calculations, necessitating approximations or the combination of categories following guidance in the Cochrane Handbook [45].

Finally, while statistically significant empathy differences were identified, their clinical relevance – such as direct impacts on patient outcomes – remains uncertain. However, the magnitude of these differences (approximately 20% absolute difference) suggests potential clinical relevance, warranting further investigation into how enhanced AI empathy might influence healthcare outcomes.

### 6.3 Recommendations for future research and practice

Future research should explore leveraging AI’s empathic advantages in text-based communication without compromising accuracy or safety. One promising approach is using AI to draft patient-facing messages, such as responses to medical or administrative queries, freeing clinician time and enhancing care quality [10]. Building on this, we propose a collaborative human–AI interaction model: clinicians produce an initial response, while AI augments these drafts by refining tone and integrating empathy, functioning as an “empathic enhancer”. This approach, ensuring accuracy through clinician oversight (as advocated by Reynolds, et al. [29] and Chen, et al. [20]), mitigates risks of AI-generated inaccuracies by having clinicians generate the core content, with AI solely providing refinement. Rigorous randomised trials are recommended to evaluate impacts on patient satisfaction and clinician workload.

Emerging voice-enabled chatbots (e.g., ChatGPT’s Advanced Voice Mode) claim capabilities to ‘respond with emotion’ and ‘pick up on non-verbal cues’ [46], yet no studies in our review compared these systems to HCPs. Given the observed empathic advantage of AI in text-only interactions, trials in telephone consultations (which account for 26% of all GP appointments [47]) could test whether this persists in voice communications.

Nearly all studies blinded raters to the source of each response (AI vs. human) to mitigate bias. However, real-world standards require disclosing AI involvement. This disclosure might have diminished perceived empathy once evaluators know the response is AI-generated.

Future studies should investigate empathy ratings in scenarios where participants/reviewers are explicitly informed if they are communicating with AI, to ascertain whether the favourable impressions seen under blinded conditions persist. Supporting this concern, Perry et al. [48] found that AI-assisted replies were initially rated more empathic than human ones in online emotional-support chats, but this advantage disappeared once users learned the responses came from AI.

Further research is necessary on how prompt design influences empathic outcomes. For instance, two studies highlighted that restricting AI’s response length – matching typical clinician brevity – reduces perceived empathy, while allowing longer responses correlates with higher ratings [10, 32]. Additionally, studies did not instruct the chatbot to emphasise empathy, potentially influencing the outcomes, with the study investigating Med-PaLM 2 suggesting it fell short in comparison to humans, as the ‘agent was not explicitly prompted to produce empathic language’ [24]. Optimising prompts to balance accuracy, brevity, and empathy is essential for real-world implementation. Beyond prompt engineering, empathy benchmarking should expand to diverse models beyond the GPT family (e.g., Claude, Llama) to identify model-specific strengths across clinical contexts.

Finally, further research should assess patients’ perceptions of empathy based on their own experiences of healthcare consultations.

## 7. Conclusion

Our review indicates that generative AI chatbots – particularly GPT-4 – are often perceived as more empathic than human practitioners in text-based interactions, a finding consistent across various clinical contexts though with notable exceptions in dermatology. While methodological limitations, such as reliance on unvalidated scales and text-only evaluations, temper these results, the compelling evidence challenges longstanding assumptions about human clinicians’ exclusive capacity for empathic communication – including assertions from the 2019 Topol Review (a UK government-commissioned roadmap for healthcare technology), which deemed ‘empathy and compassion’ an ‘essential human skill[s] that AI cannot replicate’ [8]. Future research should extend to evaluations using voice-based interactions and direct patient feedback, and ensure rigorous validation and transparency through randomised trials to uphold clinical reliability.

### Other Information

#### Protocol and registration

The protocol was developed following the Preferred Reporting Items for Systematic Reviews and Meta-Analysis Protocols (PRISMA-P) guidelines and was registered prospectively with the Open Science Framework (OSF) on 28 October 2024 (https://osf.io/2rt3f). Contrary to the original protocol’s description of a scoping review, this study was conducted as a systematic review with meta-analysis. Following protocol registration, clarifications were incorporated to refine the eligibility criteria. These included specifying the inclusion of studies utilising patient-generated data (e.g., emails), defining ‘AI’ as Large Language Models (LLMs) to improve scope precision, and broadening ‘healthcare settings’ to encompass informal health-related online contexts. In response to the observed quantitative outcome measures across included studies, the analysis plan was adapted from the originally proposed narrative synthesis to a meta-analysis, enabling the estimation of an overall effect size.

#### Contributors

AH conceived and designed the study, developed the search strategy, conducted data extraction for all records, and wrote the manuscript. AH performed full screening of all records (100% double-screened) and conducted risk of bias (RoB) assessments for all included studies. JG and AK independently screened 50% of records each and independently extracted data from 50% of records each. AK independently double-assessed RoB for all studies. JH and AB-W provided methodological input and feedback on drafts. SG reviewed a late-stage draft of the manuscript and provided feedback. All authors reviewed and approved the final manuscript. AH is the guarantor.

## Supporting information

Multimedia Appendix A (Search queries and number of results for each database)

Multimedia Appendix B Table of Full-Texts Exlcude (PICO)

Multimedia Appendix C Narrative Syntheses

Multimedia Appendix D Risk of Bias Assessment

Multimedia Appendix E Study-Specific Results

## Data Availability

All data referred to in this manuscript were obtained from previously published studies and are available in the public domain.

## Acknowledgements

We thank Selina Lock, Research Librarian, for verifying the search strategy.

## CRediT authorship contribution statement

**Alastair Howcroft:** Conceptualisation, Methodology, Software, Validation, Formal analysis, Investigation, Data curation, Writing – original draft, Writing – review & editing, Visualisation, Project administration. **Ahmad Khan:** Data curation. **Joseff Griffiths:** Data curation. **Amber Bennett-Weston:** Supervision, Writing – review & editing. **Jeremy Howick:** Supervision, Writing – review & editing. **Simon Gay:** Writing – review & editing.

---

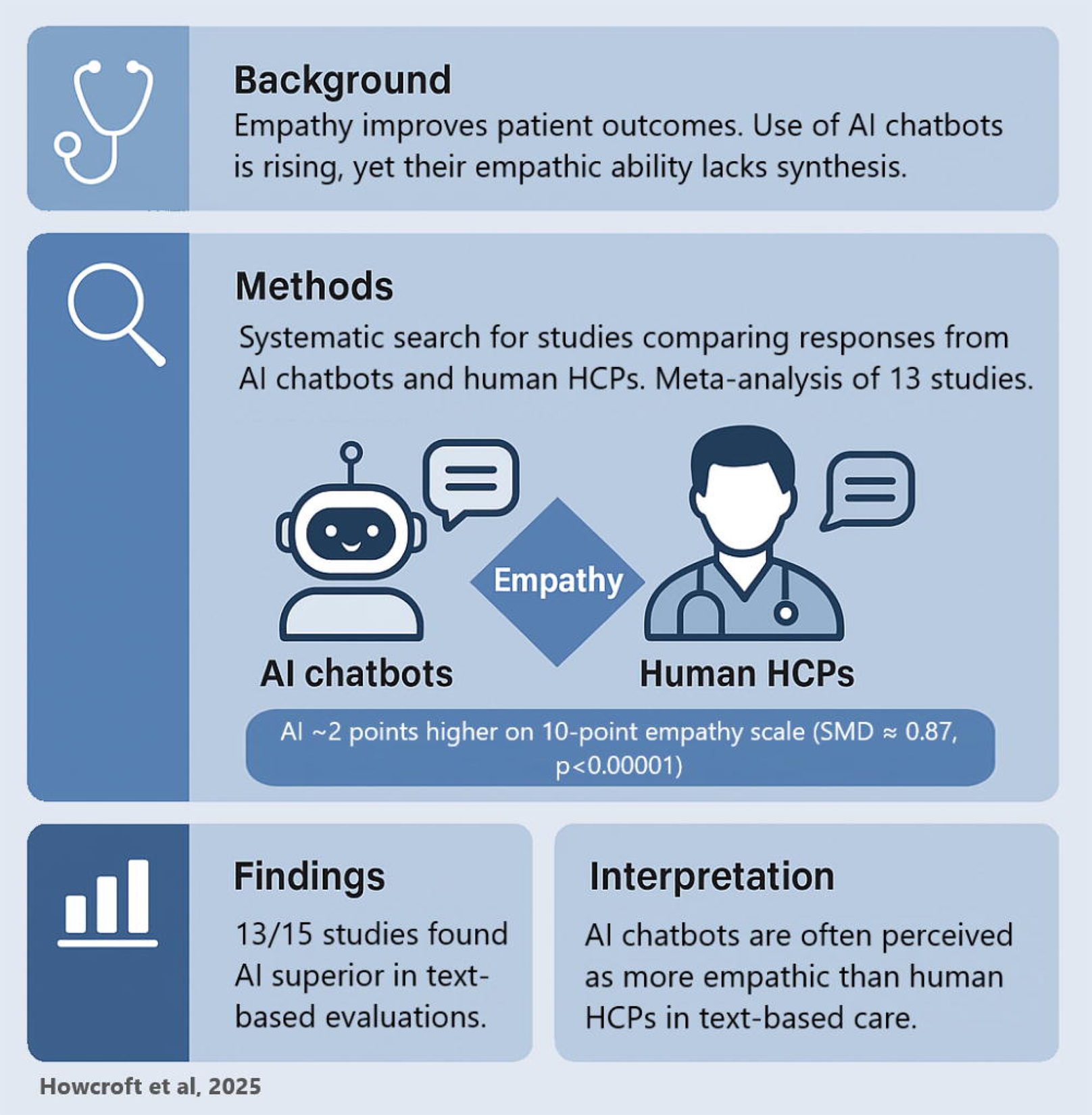

