## Supplementary material for "AI Chatbots Versus Human Healthcare Professionals: A Systematic Review and Meta-Analysis of Empathy in Patient Care": Multimedia Appendix A (Search queries and number of results for each database)

| **Database** | **Full Search String** | **Retrieved** |
| --- | --- | --- |
| IEEE Xplore | (("empath*" OR "compassion*") AND ( "chatbot" OR "chatbots" OR "Chat bot" OR "Chat bots" OR "Chat-bot" OR "Chat-bots" OR "carebot" OR "carebots" OR "Care bot" OR "Care bots" OR "Care-bot" OR "Care-bots" OR "Speech recognition software" OR "Conversational agent" OR "Conversational agents" OR "Embodied conversational agent" OR "Embodied conversational agents" OR "avatar" OR "avatars" OR "Dialog system" OR "Dialog systems" OR "Voice recognition software" OR "Virtual assistant" OR "Virtual assistants" OR "Virtual nurse" OR "Virtual nurses" OR "Virtual patient" OR "Virtual patients" OR "Virtual coach" OR "Virtual coaches" OR "Virtual agent" OR "Virtual agents" OR "Relation agent" OR "Relation agents" OR "Assistance technology" OR "Assistance technologies" OR "Intelligent assistant" OR "Intelligent assistants" OR "Digital assistant" OR "Digital assistants" OR "Natural language interface" OR "Interactive computer agent" OR "Interactive computer agents" OR "Computer-assisted instruction" OR "Natural language communication" OR "Natural language understanding" OR "Unconstrained natural language processing" OR "Artificial Intelligence" OR "ai" OR "Machine Learning" OR "Intelligent System" OR "Intelligent Systems" OR "Smart Technology" OR "Smart Technologies" OR "Cognitive Computing" OR "Neural Network" OR "Neural Networks" OR "Virtual Health Assistant" OR "Virtual Health Assistants" OR "Digital Companion" OR "Digital Companions" OR "Automated Care Provider" OR "Automated Care Providers" OR "Affective Computing System" OR "Affective Computing Systems" OR "robot" OR "robots" OR "Dialogue System" OR "Dialogue Systems" OR "Voice Assistant" OR "Voice Assistants" OR "Speech-based Agent" OR "Speech-based Agents" OR "Text-based Agent" OR "Text-based Agents" OR "Embodied Agent" OR "Embodied Agents" ) AND ( "Health Provider" OR "Health Providers" OR "Healthcare Worker" OR "Healthcare Workers" OR "Medical Practitioner" OR "Medical Practitioners" OR "Health Professional" OR "Health Professionals" OR "Medical Provider" OR "Medical Providers" OR "Medical Personnel" OR "Medical Staff" OR "Healthcare Personnel" OR "Health Practitioner" OR "Health Practitioners" OR "Medical Worker" OR "Medical Workers" OR "doctor" OR "doctors" OR "physician" OR "physicians" OR "General Practitioner" OR "General Practitioners" OR "gp" OR "gps" OR "clinician" OR "clinicians" OR "nurse*" OR "midwif*" OR "Physician Assistant" OR "Physician Assistants" OR "Medical Assistant" OR "Medical Assistants" OR "paramedic" OR "paramedics" OR "Emergency Medical Technician" OR "Emergency Medical Technicians" OR "Home Health Aide" OR "Home Health Aides" OR "psychiatrist" OR "psychiatrists" OR "psychologist" OR "psychologists" OR "therapist" OR "therapists" OR "counselor" OR "counselors" OR "counsellor" OR "counsellors" OR "Social Worker" OR "Social Workers" OR "dietitian" OR "dietitians" OR "nutritionist" OR "nutritionists" OR "pharmacist" OR "pharmacists" OR "dentist" OR "dentists" OR "surgeon" OR "surgeons" OR "cardiologist" OR "cardiologists" OR "oncologist" OR "oncologists" OR "radiologist" OR "radiologists" OR "anesthesiologist" OR "anesthesiologists" OR "pediatrician" OR "pediatricians" OR "geriatrician" OR "geriatricians" OR "chiropractor" OR "chiropractors" OR "optometrist" OR "optometrists" OR "Speech-Language Pathologist" OR "Speech-Language Pathologists" OR "audiologist" OR "audiologists" OR "ophthalmologist" OR "ophthalmologists" OR "Medical Laboratory Technician" OR "Medical Laboratory Technicians" OR "Medical Technologist" OR "Medical Technologists" OR "sonographer" OR "sonographers" OR "technologist" OR "technologists" OR "interpreter" OR "interpreters" OR "transcriptionist" OR "transcriptionists" OR "technician" OR "technicians" OR "Health Information Technician" OR "Health Information Technicians" OR "acupuncturist" OR "acupuncturists" OR "homeopath" OR "homeopaths" OR "naturopath" OR "naturopaths" OR "Health Worker" OR "Health Workers" OR "kinesiologist" OR "kinesiologists" OR "veterinarian" OR "veterinarians" OR "Care worker" OR "Care workers" OR "caregiver" OR "caregivers" OR "Care giver" OR "Care givers" ) ) | 148 |
| MEDLINE (PubMed) | ("Empathy"[Mesh] OR "Compassion"[Mesh] OR  empathy[tiab] OR empathic[tiab] OR empathetic[tiab] OR empath[tiab] OR empathically[tiab] OR  compassion[tiab] OR compassionate[tiab] OR compassionately[tiab])  AND  (  "Artificial Intelligence"[Mesh] OR "Robotics"[Mesh] OR "Speech Recognition Software"[Mesh] OR  "Natural Language Processing"[Mesh] OR "Machine Learning"[Mesh] OR "Virtual Reality"[Mesh] OR  "Computer-Assisted Instruction"[Mesh] OR "User-Computer Interface"[Mesh] OR  "Chatbot"[tiab] OR "Chatbots"[tiab] OR  "Carebot"[tiab] OR "Carebots"[tiab] OR ("Care"[tiab] AND "bot"[tiab]) OR ("Care"[tiab] AND "bots"[tiab]) OR  "Speech recognition software"[tiab] OR  "Conversational agent"[tiab] OR "Conversational agents"[tiab] OR  "Embodied conversational agent"[tiab] OR "Embodied conversational agents"[tiab] OR  avatar[tiab] OR avatars[tiab] OR  "Dialog system"[tiab] OR "Dialog systems"[tiab] OR  "Voice recognition software"[tiab] OR  "Virtual assistant"[tiab] OR "Virtual assistants"[tiab] OR  "Virtual nurse"[tiab] OR "Virtual nurses"[tiab] OR  "Virtual patient"[tiab] OR "Virtual patients"[tiab] OR  "Virtual coach"[tiab] OR "Virtual coaches"[tiab] OR  "Virtual agent"[tiab] OR "Virtual agents"[tiab] OR  "Relational agent"[tiab] OR "Relational agents"[tiab] OR  ("Assistance"[tiab] AND "technology"[tiab]) OR ("Assistance"[tiab] AND "technologies"[tiab]) OR  ("Interactive"[tiab] AND "computer"[tiab] AND "agent"[tiab]) OR ("Interactive"[tiab] AND "computer"[tiab] AND "agents"[tiab]) OR  "Natural language interface"[tiab] OR  ("Natural"[tiab] AND "language"[tiab] AND "communication"[tiab]) OR  "Unconstrained natural language processing"[tiab] OR  "Artificial intelligence"[tiab] OR "AI"[tiab] OR  "Machine learning"[tiab] OR  "Intelligent system"[tiab] OR "Intelligent systems"[tiab] OR  "Smart technology"[tiab] OR "Smart technologies"[tiab] OR  "Cognitive computing"[tiab] OR  "Neural network"[tiab] OR "Neural networks"[tiab] OR  ("Virtual health"[tiab] AND "assistant"[tiab]) OR ("Virtual health"[tiab] AND "assistants"[tiab]) OR  "Digital companion"[tiab] OR "Digital companions"[tiab] OR  ("Automated care"[tiab] AND "provider"[tiab]) OR ("Automated care"[tiab] AND "providers"[tiab]) OR  "Affective computing"[tiab] OR ("Affective computing"[tiab] AND "system"[tiab]) OR ("Affective computing"[tiab] AND "systems"[tiab]) OR  robot[tiab] OR robots[tiab] OR  "Dialogue system"[tiab] OR "Dialogue systems"[tiab] OR  "Voice assistant"[tiab] OR "Voice assistants"[tiab] OR  ("Speech-based"[tiab] AND "agent"[tiab]) OR ("Speech-based"[tiab] AND "agents"[tiab]) OR  ("Text-based"[tiab] AND "agent"[tiab]) OR ("Text-based"[tiab] AND "agents"[tiab]) OR  "Embodied agent"[tiab] OR "Embodied agents"[tiab]  )  AND  (  "Health Personnel"[Mesh] OR "Physicians"[Mesh] OR "Nurses"[Mesh] OR "Medical Staff"[Mesh] OR  "Allied Health Personnel"[Mesh] OR "Health Occupations"[Mesh] OR  doctor[tiab] OR doctors[tiab] OR  physician[tiab] OR physicians[tiab] OR  "General practitioner"[tiab] OR "General practitioners"[tiab] OR  gp[tiab] OR gps[tiab] OR  clinician[tiab] OR clinicians[tiab] OR  nurse[tiab] OR nurses[tiab] OR  midwife[tiab] OR midwives[tiab] OR  "Physician assistant"[tiab] OR "Physician assistants"[tiab] OR  "Medical assistant"[tiab] OR "Medical assistants"[tiab] OR  paramedic[tiab] OR paramedics[tiab] OR  "Emergency medical technician"[tiab] OR "Emergency medical technicians"[tiab] OR  "Home health aide"[tiab] OR "Home health aides"[tiab] OR  psychiatrist[tiab] OR psychiatrists[tiab] OR  psychologist[tiab] OR psychologists[tiab] OR  therapist[tiab] OR therapists[tiab] OR  counselor[tiab] OR counselors[tiab] OR  counsellor[tiab] OR counsellors[tiab] OR  "Social worker"[tiab] OR "Social workers"[tiab] OR  dietitian[tiab] OR dietitians[tiab] OR  nutritionist[tiab] OR nutritionists[tiab] OR  pharmacist[tiab] OR pharmacists[tiab] OR  dentist[tiab] OR dentists[tiab] OR  surgeon[tiab] OR surgeons[tiab] OR  cardiologist[tiab] OR cardiologists[tiab] OR  oncologist[tiab] OR oncologists[tiab] OR  radiologist[tiab] OR radiologists[tiab] OR  anesthesiologist[tiab] OR anesthesiologists[tiab] OR  pediatrician[tiab] OR pediatricians[tiab] OR  geriatrician[tiab] OR geriatricians[tiab] OR  chiropractor[tiab] OR chiropractors[tiab] OR  optometrist[tiab] OR optometrists[tiab] OR  "Speech-language pathologist"[tiab] OR "Speech-language pathologists"[tiab] OR  audiologist[tiab] OR audiologists[tiab] OR  ophthalmologist[tiab] OR ophthalmologists[tiab] OR  "Medical laboratory technician"[tiab] OR "Medical laboratory technicians"[tiab] OR  "Medical technologist"[tiab] OR "Medical technologists"[tiab] OR  sonographer[tiab] OR sonographers[tiab] OR  technologist[tiab] OR technologists[tiab] OR  interpreter[tiab] OR interpreters[tiab] OR  transcriptionist[tiab] OR transcriptionists[tiab] OR  technician[tiab] OR technicians[tiab] OR  "Health information technician"[tiab] OR "Health information technicians"[tiab] OR  acupuncturist[tiab] OR acupuncturists[tiab] OR  homeopath[tiab] OR homeopaths[tiab] OR  naturopath[tiab] OR naturopaths[tiab] OR  "Health worker"[tiab] OR "Health workers"[tiab] OR  kinesiologist[tiab] OR kinesiologists[tiab] OR  veterinarian[tiab] OR veterinarians[tiab] OR  "Care worker"[tiab] OR "Care workers"[tiab] OR  caregiver[tiab] OR caregivers[tiab] OR  "Care giver"[tiab] OR "Care givers"[tiab]  ) | 358 |
| CINAHL | (  TI (empath* OR compassion*) OR  AB (empath* OR compassion*)  )  AND  (  TI (  chatbot* OR "Chat bot*" OR "Chat-bot*" OR  carebot* OR "Care bot*" OR "Care-bot*" OR  "Speech recognition software" OR  "Conversational agent*" OR  "Embodied conversational agent*" OR  avatar* OR  "Dialog system*" OR  "Voice recognition software" OR  "Virtual assistan*" OR  "Virtual nurs*" OR  "Virtual patient*" OR  "Virtual coach*" OR  "Virtual agent*" OR  "Relation agent*" OR  "Assistance technol*" OR  "Intelligent assistan*" OR  "Digital assistan*" OR  "Natural language interface" OR  "Interactive computer agent*" OR  "Computer-assisted instruction" OR  "Natural language communication" OR  "Natural language understanding" OR  "Unconstrained natural language processing" OR  "Artificial Intelligence*" OR  ai OR  "Machine Learning" OR  "Intelligent System*" OR  "Smart Technology*" OR  "Cognitive Computing*" OR  "Neural Network*" OR  "Virtual Health Assistant*" OR  "Digital Companion*" OR  "Automated Care Provider*" OR  "Affective Computing System*" OR  robot* OR  "Virtual Assistant*" OR  "Dialogue System*" OR  "Voice Assistant*" OR  "Speech-based Agent*" OR  "Text-based Agent*" OR  "Embodied Agent*"  ) OR  AB (  chatbot* OR "Chat bot*" OR "Chat-bot*" OR  carebot* OR "Care bot*" OR "Care-bot*" OR  "Speech recognition software" OR  "Conversational agent*" OR  "Embodied conversational agent*" OR  avatar* OR  "Dialog system*" OR  "Voice recognition software" OR  "Virtual assistan*" OR  "Virtual nurs*" OR  "Virtual patient*" OR  "Virtual coach*" OR  "Virtual agent*" OR  "Relation agent*" OR  "Assistance technol*" OR  "Intelligent assistan*" OR  "Digital assistan*" OR  "Natural language interface" OR  "Interactive computer agent*" OR  "Computer-assisted instruction" OR  "Natural language communication" OR  "Natural language understanding" OR  "Unconstrained natural language processing" OR  "Artificial Intelligence*" OR  ai OR  "Machine Learning" OR  "Intelligent System*" OR  "Smart Technology*" OR  "Cognitive Computing*" OR  "Neural Network*" OR  "Virtual Health Assistant*" OR  "Digital Companion*" OR  "Automated Care Provider*" OR  "Affective Computing System*" OR  robot* OR  "Virtual Assistant*" OR  "Dialogue System*" OR  "Voice Assistant*" OR  "Speech-based Agent*" OR  "Text-based Agent*" OR  "Embodied Agent*"  )  )  AND  (  TI (  "Health Provider*" OR "Healthcare Worker*" OR  "Medical Practitioner*" OR "Health Professional*" OR  "Medical Provider*" OR "Medical Personnel*" OR  "Medical Staff*" OR "Healthcare Personnel*" OR  "Health Practitioner*" OR "Medical Worker*" OR  doctor* OR physician* OR  "General Practitioner*" OR gp OR gps OR  clinician* OR nurse* OR midwif* OR  "Physician Assistant*" OR "Medical Assistant*" OR  paramedic* OR "Emergency Medical Technician*" OR  "Home Health Aide*" OR psychiatrist* OR  psychologist* OR therapist* OR  counselor* OR counsellor* OR  "Social Worker*" OR dietitian* OR  nutritionist* OR pharmacist* OR  dentist* OR surgeon* OR  cardiologist* OR oncologist* OR  radiologist* OR anesthesiologist* OR  pediatrician* OR geriatrician* OR  chiropractor* OR optometrist* OR  "Speech-Language Pathologist*" OR audiologist* OR  ophthalmologist* OR "Medical Laboratory Technician*" OR  "Medical Technologist*" OR sonographer* OR  technologist* OR interpreter* OR  transcriptionist* OR technician* OR  "Health Information Technician*" OR  acupuncturist* OR homeopath* OR  naturopath* OR "Health Worker*" OR  kinesiologist* OR veterinarian* OR  "Care worker*" OR caregiver* OR "Care giver*"  ) OR  AB (  "Health Provider*" OR "Healthcare Worker*" OR  "Medical Practitioner*" OR "Health Professional*" OR  "Medical Provider*" OR "Medical Personnel*" OR  "Medical Staff*" OR "Healthcare Personnel*" OR  "Health Practitioner*" OR "Medical Worker*" OR  doctor* OR physician* OR  "General Practitioner*" OR gp OR gps OR  clinician* OR nurse* OR midwif* OR  "Physician Assistant*" OR "Medical Assistant*" OR  paramedic* OR "Emergency Medical Technician*" OR  "Home Health Aide*" OR psychiatrist* OR  psychologist* OR therapist* OR  counselor* OR counsellor* OR  "Social Worker*" OR dietitian* OR  nutritionist* OR pharmacist* OR  dentist* OR surgeon* OR  cardiologist* OR oncologist* OR  radiologist* OR anesthesiologist* OR  pediatrician* OR geriatrician* OR  chiropractor* OR optometrist* OR  "Speech-Language Pathologist*" OR audiologist* OR  ophthalmologist* OR "Medical Laboratory Technician*" OR  "Medical Technologist*" OR sonographer* OR  technologist* OR interpreter* OR  transcriptionist* OR technician* OR  "Health Information Technician*" OR  acupuncturist* OR homeopath* OR  naturopath* OR "Health Worker*" OR  kinesiologist* OR veterinarian* OR  "Care worker*" OR caregiver* OR "Care giver*"  )  ) | 99 |
| PsycINFO | (  TI (empath* OR compassion*) OR  AB (empath* OR compassion*) OR  KW (empath* OR compassion*)  )  AND  (  TI (chatbot* OR "chat bot*" OR "chat-bot*" OR carebot* OR "care bot*" OR "care-bot*" OR "speech recognition software" OR "conversational agent*" OR "embodied conversational agent*" OR avatar* OR "dialog system*" OR "voice recognition software" OR "virtual assistan*" OR "virtual nurs*" OR "virtual patient*" OR "virtual coach*" OR "virtual agent*" OR "relation agent*" OR "assistance technol*" OR "intelligent assistan*" OR "digital assistan*" OR "natural language interface" OR "interactive computer agent*" OR "computer-assisted instruction" OR "natural language communication" OR "natural language understanding" OR "unconstrained natural language processing" OR "artificial intelligence*" OR ai OR "machine learning" OR "intelligent system*" OR "smart technology*" OR "cognitive computing*" OR "neural network*" OR "virtual health assistant*" OR "digital companion*" OR "automated care provider*" OR "affective computing system*" OR robot* OR "virtual assistant*" OR "dialogue system*" OR "voice assistant*" OR "speech-based agent*" OR "text-based agent*" OR "embodied agent*") OR  AB (chatbot* OR "chat bot*" OR "chat-bot*" OR carebot* OR "care bot*" OR "care-bot*" OR "speech recognition software" OR "conversational agent*" OR "embodied conversational agent*" OR avatar* OR "dialog system*" OR "voice recognition software" OR "virtual assistan*" OR "virtual nurs*" OR "virtual patient*" OR "virtual coach*" OR "virtual agent*" OR "relation agent*" OR "assistance technol*" OR "intelligent assistan*" OR "digital assistan*" OR "natural language interface" OR "interactive computer agent*" OR "computer-assisted instruction" OR "natural language communication" OR "natural language understanding" OR "unconstrained natural language processing" OR "artificial intelligence*" OR ai OR "machine learning" OR "intelligent system*" OR "smart technology*" OR "cognitive computing*" OR "neural network*" OR "virtual health assistant*" OR "digital companion*" OR "automated care provider*" OR "affective computing system*" OR robot* OR "virtual assistant*" OR "dialogue system*" OR "voice assistant*" OR "speech-based agent*" OR "text-based agent*" OR "embodied agent*") OR  KW (chatbot* OR "chat bot*" OR "chat-bot*" OR carebot* OR "care bot*" OR "care-bot*" OR "speech recognition software" OR "conversational agent*" OR "embodied conversational agent*" OR avatar* OR "dialog system*" OR "voice recognition software" OR "virtual assistan*" OR "virtual nurs*" OR "virtual patient*" OR "virtual coach*" OR "virtual agent*" OR "relation agent*" OR "assistance technol*" OR "intelligent assistan*" OR "digital assistan*" OR "natural language interface" OR "interactive computer agent*" OR "computer-assisted instruction" OR "natural language communication" OR "natural language understanding" OR "unconstrained natural language processing" OR "artificial intelligence*" OR ai OR "machine learning" OR "intelligent system*" OR "smart technology*" OR "cognitive computing*" OR "neural network*" OR "virtual health assistant*" OR "digital companion*" OR "automated care provider*" OR "affective computing system*" OR robot* OR "virtual assistant*" OR "dialogue system*" OR "voice assistant*" OR "speech-based agent*" OR "text-based agent*" OR "embodied agent*")  )  AND  (  TI ("health provider*" OR "healthcare worker*" OR "medical practitioner*" OR "health professional*" OR "medical provider*" OR "medical personnel*" OR "medical staff*" OR "healthcare personnel*" OR "health practitioner*" OR "medical worker*" OR doctor* OR physician* OR "general practitioner*" OR gp OR gps OR clinician* OR nurse* OR midwif* OR "physician assistant*" OR "medical assistant*" OR paramedic* OR "emergency medical technician*" OR "home health aide*" OR psychiatrist* OR psychologist* OR therapist* OR counselor* OR counsellor* OR "social worker*" OR dietitian* OR nutritionist* OR pharmacist* OR dentist* OR surgeon* OR cardiologist* OR oncologist* OR radiologist* OR anesthesiologist* OR pediatrician* OR geriatrician* OR chiropractor* OR optometrist* OR "speech-language pathologist*" OR audiologist* OR ophthalmologist* OR "medical laboratory technician*" OR "medical technologist*" OR sonographer* OR technologist* OR interpreter* OR transcriptionist* OR technician* OR "health information technician*" OR acupuncturist* OR homeopath* OR naturopath* OR "health worker*" OR kinesiologist* OR veterinarian* OR "care worker*" OR caregiver* OR "care giver*") OR  AB ("health provider*" OR "healthcare worker*" OR "medical practitioner*" OR "health professional*" OR "medical provider*" OR "medical personnel*" OR "medical staff*" OR "healthcare personnel*" OR "health practitioner*" OR "medical worker*" OR doctor* OR physician* OR "general practitioner*" OR gp OR gps OR clinician* OR nurse* OR midwif* OR "physician assistant*" OR "medical assistant*" OR paramedic* OR "emergency medical technician*" OR "home health aide*" OR psychiatrist* OR psychologist* OR therapist* OR counselor* OR counsellor* OR "social worker*" OR dietitian* OR nutritionist* OR pharmacist* OR dentist* OR surgeon* OR cardiologist* OR oncologist* OR radiologist* OR anesthesiologist* OR pediatrician* OR geriatrician* OR chiropractor* OR optometrist* OR "speech-language pathologist*" OR audiologist* OR ophthalmologist* OR "medical laboratory technician*" OR "medical technologist*" OR sonographer* OR technologist* OR interpreter* OR transcriptionist* OR technician* OR "health information technician*" OR acupuncturist* OR homeopath* OR naturopath* OR "health worker*" OR kinesiologist* OR veterinarian* OR "care worker*" OR caregiver* OR "care giver*") OR  KW ("health provider*" OR "healthcare worker*" OR "medical practitioner*" OR "health professional*" OR "medical provider*" OR "medical personnel*" OR "medical staff*" OR "healthcare personnel*" OR "health practitioner*" OR "medical worker*" OR doctor* OR physician* OR "general practitioner*" OR gp OR gps OR clinician* OR nurse* OR midwif* OR "physician assistant*" OR "medical assistant*" OR paramedic* OR "emergency medical technician*" OR "home health aide*" OR psychiatrist* OR psychologist* OR therapist* OR counselor* OR counsellor* OR "social worker*" OR dietitian* OR nutritionist* OR pharmacist* OR dentist* OR surgeon* OR cardiologist* OR oncologist* OR radiologist* OR anesthesiologist* OR pediatrician* OR geriatrician* OR chiropractor* OR optometrist* OR "speech-language pathologist*" OR audiologist* OR ophthalmologist* OR "medical laboratory technician*" OR "medical technologist*" OR sonographer* OR technologist* OR interpreter* OR transcriptionist* OR technician* OR "health information technician*" OR acupuncturist* OR homeopath* OR naturopath* OR "health worker*" OR kinesiologist* OR veterinarian* OR "care worker*" OR caregiver* OR "care giver*")  ) | 116 |
| Scopus | (TITLE-ABS-KEY("Artificial Intelligence" OR AI OR "Machine Learning" OR "Intelligent System" OR "Smart Technology" OR "Cognitive Computing" OR "Neural Network" OR "Chatbot" OR "Care-bot" OR "Conversational Agent" OR "Digital Assistant" OR "Virtual Health Assistant" OR "Digital Companion" OR "Health Chatbot" OR "Interactive Agent" OR "Automated Care Provider" OR "Robotic Companion" OR "Social Robot" OR "Socially Assistive Robot" OR "Human-Robot Interaction" OR "Empathic Robot" OR "Affective Computing System" OR "Robotic Healthcare Provider" OR "Virtual Assistant" OR "Dialogue System" OR "Voice Assistant" OR "Speech-based Agent" OR "Text-based Agent" OR "Embodied Agent"))  AND  (TITLE-ABS-KEY("Empathy" OR "Empathic" OR "Empathetic"))  AND  (TITLE-ABS-KEY("Healthcare" OR "Patient Care" OR "Medical Care" OR "Health Services" OR "Healthcare Delivery" OR "Clinical Care" OR "Clinical Interaction" OR "Healthcare Setting" OR "Medical Setting" OR "Health Provider" OR "Healthcare Worker" OR "Medical Practitioner" OR "Health Professional" OR "Medical Provider" OR "Doctor" OR "Physician" OR "Nurse" OR "Clinician" OR "Primary Care" OR "Hospital Staff" OR "Medical Personnel" OR "Caregiver" OR "Psychiatrist" OR "Psychologist" OR "Therapist" OR "Counselor" OR "Social Worker" OR "Physician Assistant" OR "Nurse Practitioner" OR "Pharmacist" OR "Dentist" OR "Radiologist" OR "Anesthesiologist" OR "Surgeon" OR "Oncologist" OR "Cardiologist" OR "Pediatrician" OR "Geriatrician" OR "Chiropractor" OR "Optometrist" OR "Speech-Language Pathologist" OR "Dietitian" OR "Nutritionist" OR "Midwife" OR "Paramedic" OR "Emergency Medical Technician")) | 653 |
| Cochrane Library | (empath* OR compassion*) AND ( chatbot* OR (chat NEXT bot*) OR carebot* OR (care NEXT bot*) OR "speech recognition software" OR (conversational NEXT agent*) OR (embodied NEXT conversational NEXT agent*) OR avatar* OR (dialog NEXT system*) OR (voice NEXT recognition NEXT software) OR (virtual NEXT assistant*) OR (virtual NEXT nurse*) OR (virtual NEXT patient*) OR (virtual NEXT coach*) OR (virtual NEXT agent*) OR (relational NEXT agent*) OR (assistance NEXT technolog*) OR (intelligent NEXT assistant*) OR (digital NEXT assistant*) OR (natural NEXT language NEXT interface) OR (interactive NEXT computer NEXT agent*) OR (computer NEXT assisted NEXT instruction) OR (natural NEXT language NEXT communication) OR (natural NEXT language NEXT understanding) OR (unconstrained NEXT natural NEXT language NEXT processing) OR (artificial NEXT intelligence) OR ai OR (machine NEXT learning) OR (intelligent NEXT system*) OR (smart NEXT technolog*) OR (cognitive NEXT computing*) OR (neural NEXT network*) OR (virtual NEXT health NEXT assistant*) OR (digital NEXT companion*) OR (automated NEXT care NEXT provider*) OR (affective NEXT computing NEXT system*) OR robot* OR (dialogue NEXT system*) OR (voice NEXT assistant*) OR (speech NEXT based NEXT agent*) OR (text NEXT based NEXT agent*) OR (embodied NEXT agent*) ) AND ( (health NEXT provider*) OR (healthcare NEXT worker*) OR (medical NEXT practitioner*) OR (health NEXT professional*) OR (medical NEXT provider*) OR (medical NEXT personnel*) OR (medical NEXT staff*) OR (healthcare NEXT personnel*) OR (health NEXT practitioner*) OR (medical NEXT worker*) OR doctor* OR physician* OR (general NEXT practitioner*) OR gp OR gps OR clinician* OR nurse* OR midwif* OR (physician NEXT assistant*) OR (medical NEXT assistant*) OR paramedic* OR (emergency NEXT medical NEXT technician*) OR (home NEXT health NEXT aide*) OR psychiatrist* OR psychologist* OR therapist* OR counselor* OR counsellor* OR (social NEXT worker*) OR dietitian* OR nutritionist* OR pharmacist* OR dentist* OR surgeon* OR cardiologist* OR oncologist* OR radiologist* OR anesthesiologist* OR pediatrician* OR geriatrician* OR chiropractor* OR optometrist* OR (speech NEXT language NEXT pathologist*) OR audiologist* OR ophthalmologist* OR (medical NEXT laboratory NEXT technician*) OR (medical NEXT technologist*) OR sonographer* OR technologist* OR interpreter* OR transcriptionist* OR technician* OR (health NEXT information NEXT technician*) OR acupuncturist* OR homeopath* OR naturopath* OR (health NEXT worker*) OR kinesiologist* OR veterinarian* OR (care NEXT worker*) OR caregiver* OR (care NEXT giver*) ) | 54 |
| ClinicalTrials.gov | ("empathy" OR "empathic" OR empathetic OR "empathize" OR "empathised" OR "empathising" OR "empathized" OR "empathise" OR "empathised" OR "empathising" OR "compassion" OR "compassionate") AND ("chatbot" OR "chat bot" OR "chat-bot" OR "carebot" OR "care bot" OR "care-bot" OR "speech recognition software" OR "conversational agent" OR "embodied conversational agent" OR "avatar" OR "dialog system" OR "dialogue system" OR "voice recognition software" OR "virtual assistant" OR "virtual nurse" OR "virtual patient" OR "virtual coach" OR "virtual agent" OR "relational agent" OR "assistance technology" OR "intelligent assistant" OR "digital assistant" OR "natural language interface" OR "interactive computer agent" OR "computer-assisted instruction" OR "natural language communication" OR "natural language understanding" OR "unconstrained natural language processing" OR "artificial intelligence" OR "AI" OR "machine learning" OR "intelligent system" OR "smart technology" OR "cognitive computing" OR "neural network" OR "virtual health assistant" OR "digital companion" OR "automated care provider" OR "affective computing system" OR "robot" OR "speech-based agent" OR "text-based agent" OR "embodied agent") AND ("health provider" OR "healthcare worker" OR "medical practitioner" OR "health professional" OR "medical provider" OR "medical personnel" OR "medical staff" OR "healthcare personnel" OR "health practitioner" OR "medical worker" OR "doctor" OR "physician" OR "general practitioner" OR "GP" OR "GPs" OR "clinician" OR "nurse" OR "midwife" OR "midwives" OR "physician assistant" OR "medical assistant" OR "paramedic" OR "emergency medical technician" OR "home health aide" OR "psychiatrist" OR "psychologist" OR "therapist" OR "counselor" OR "counsellor" OR "social worker" OR "dietitian" OR "nutritionist" OR "pharmacist" OR "dentist" OR "surgeon" OR "cardiologist" OR "oncologist" OR "radiologist" OR "anesthesiologist" OR "pediatrician" OR "geriatrician" OR "chiropractor" OR "optometrist" OR "speech-language pathologist" OR "audiologist" OR "ophthalmologist" OR "medical laboratory technician" OR "medical technologist" OR "sonographer" OR "technologist" OR "interpreter" OR "transcriptionist" OR "technician" OR "health information technician" OR "acupuncturist" OR "homeopath" OR "naturopath" OR "health worker" OR "kinesiologist" OR "veterinarian" OR "care worker" OR "caregiver" OR "care giver") | 28 |
| ICTRP | ("empathy" OR "empathic" OR "empathetic" OR "empathize" OR "empathised" OR "empathising" OR "compassion" OR "compassionate")  AND ("chatbot" OR "carebot" OR "speech recognition" OR "conversational agent" OR "embodied agent" OR "dialogue system" OR "voice recognition"  OR "virtual assistant" OR "virtual nurse" OR "virtual patient" OR "virtual coach" OR "relational agent" OR "assistance technology"  OR "intelligent assistant" OR "digital assistant" OR "natural language" OR "interactive agent" OR "computer-assisted" OR "artificial intelligence"  OR "AI" OR "machine learning" OR "intelligent system" OR "smart technology" OR "cognitive computing" OR "neural network" OR "digital companion"  OR "affective computing" OR "robot" OR "speech-based agent" OR "text-based agent")  AND ("health provider" OR "healthcare worker" OR "medical practitioner" OR "health professional" OR "doctor" OR "physician"  OR "general practitioner" OR "GP" OR "clinician" OR "nurse" OR "midwife" OR "medical assistant" OR "paramedic"  OR "emergency medical technician" OR "psychiatrist" OR "psychologist" OR "therapist" OR "counselor" OR "social worker"  OR "dietitian" OR "nutritionist" OR "pharmacist" OR "dentist" OR "surgeon" OR "radiologist" OR "pediatrician"  OR "geriatrician" OR "technician" OR "health worker" OR "caregiver") | 3 |
| ISRCTN Registry | (empathy OR empathic OR compassionate)  AND (chatbot OR "chat bot" OR carebot OR "speech recognition" OR "conversational agent" OR avatar OR "dialog system" OR "virtual assistant" OR "virtual nurse" OR "virtual agent" OR "intelligent assistant" OR "digital assistant" OR "natural language" OR "artificial intelligence" OR AI OR "machine learning" OR robot)  AND ("health provider" OR "medical practitioner" OR doctor OR physician OR clinician OR nurse OR therapist OR counselor OR "social worker" OR pharmacist OR dentist OR surgeon OR psychiatrist OR psychologist OR technician OR caregiver) | 2 |

**Grey Literature Search**

In addition to database searches, Google Scholar was used to identify grey literature. The first five pages of search results for the following search query were reviewed, as recommended by the Joanna Briggs Institute (JBI):

Search query:

("empathy" OR "empathic" OR "empathetic" OR "empathize" OR "empathised" OR "empathising" OR "compassion" OR "compassionate") AND ("chatbot" OR "chat bot" OR "carebot" OR "care bot" OR "speech recognition" OR "conversational agent" OR "virtual assistant" OR "digital assistant" OR "AI" OR "artificial intelligence" OR "machine learning" OR "neural network") AND ("healthcare worker" OR "medical practitioner" OR "health provider" OR "doctor" OR "physician" OR "nurse" OR "clinician" OR "therapist" OR "caregiver" OR "health professional")

Date searched: 12/11/2024.

Three additional relevant studies, not already identified in the database or register searches, were retrieved from Google Scholar.
