## Supplementary material for "AI Chatbots Versus Human Healthcare Professionals: A Systematic Review and Meta-Analysis of Empathy in Patient Care": Multimedia Appendix B Table of Full-Texts Exlcude (PICO)

### Multimedia Appendix B. Studies excluded during full-text screening

| **No** | **First author & year** | **Article title** | **Reason for exclusion** |
| --- | --- | --- | --- |
| 1 | Holeva et al., 2022 | Social robots as tools in special education (SRTSE) | Study design |
| 2 | Broadbent, 2024 | Can AI deliver empathetic medical consultations? | Ongoing trial |
| 3 | Ahmed et al., 2020 | Robo-Friend: Can a Social Robot Empathize with Your Feelings Effectively? | Comparison |
| 4 | Barnett et al., 2021 | Enacting ‘more-than-human’ care: Clients’ and counsellors’ views on the multiple affordances of chatbots in alcohol and other drug counselling | Intervention & comparison |
| 5 | Bickmore et al., 2010 | Relational agents in clinical psychiatry | Comparison |
| 6 | Bickmore et al., 2010 | Response to a relational agent by hospital patients with depressive symptoms | Comparison |
| 7 | Bickmore et al., 2009 | Taking the time to care: Empowering low health literacy hospital patients with virtual nurse agents | Study design |
| 8 | Daher et al., 2023 | Breaking barriers: can ChatGPT compete with a shoulder and elbow specialist in diagnosis and management? | Outcome |
| 9 | Dino et al., 2022 | Nursing and human-computer interaction in healthcare robots for older people: An integrative review | Outcome |
| 10 | Do et al., 2016 | Empathic Virual Assistant for Healthcare Information with Positive Emotional Experience | Study design |
| 11 | Durairaj et al., 2024 | Artificial Intelligence Versus Expert Plastic Surgeon: Comparative Study Shows ChatGPT "Wins" Rhinoplasty Consultations: Should We Be Worried? | Participants |
| 12 | Espinoza et al., 2023 | Supporting dementia caregivers in Peru through chatbots: generative AI vs structured conversations | Study design |
| 13 | Ibrahim et al., 2024 | Utilizing Support Vector Machine to Help with Social Robot Therapy for Autism Spectrum Disorder | Outcome |
| 14 | Weng, 2024 | AI-powered Chatbot vs. Human Counselors in Smoking Cessation | Ongoing trial |
| 15 | Wang, 2024 | RCT Supportive Care Outpatient RIC HCT | Ongoing trial |
| 16 | Pontier et al., 2008 | A virtual therapist that responds empathically to your answers | Comparison |
| 17 | Zhang et al., 2023 | Consumers' responses to personalized service from medical artificial intelligence and human doctors | Participants |
| 18 | Karhiy et al., 2024 | Can a virtual human increase mindfulness and reduce stress? A randomised trial | Intervention |
| 19 | Chen et al., 2024 | Evaluating the potential of GPT-4 in assisting communication in clinical anaesthesia | Comparison |
