## Supplementary material for "AI Chatbots Versus Human Healthcare Professionals: A Systematic Review and Meta-Analysis of Empathy in Patient Care": Multimedia Appendix C Narrative Syntheses

### Multimedia Appendix C. Narrative Synthesis of the 15 Included Studies

*Subgroup: GPT-3.5 Narrative Synthesis*

Maida (2024) investigated responses to four common neurologic inquiries emailed by multiple sclerosis patients, reporting that GPT-3.5 achieved a 1.38-point lead on the (10-50) CARE scale (95% CI: 0.65–2.11), equivalent to a 3–4% higher mean empathy rating than neurologists, with a SMD of 0.15 (95% CI: 0.07–0.23). The following three studies utilised a 1-5 Likert Scale. Wan (2024) used a supervised GPT-3.5 system called “SSPEC” in outpatient reception settings, finding that it significantly outperformed a nurse-only reception model (mean empathy 4.14 ± 0.98 vs. 3.27 ± 1.22; p < 0.001) with a SMD of 0.79 (95% CI: 0.70–0.87), as evaluated by three receptionist nurses and three non-experts. Ayers (2023) provided one the most striking instances of AI exceeding humans (SMD = 1.91; 95% CI: 1.67 – 2.15), using actual patient queries from a public social media forum. They found that HCPs preferred GPT-3.5’s responses 78.6% of the time (95% CI: 75%–81.8%); notably, 45.1% of chatbot replies were considered “empathetic” or “very empathetic,” compared to just 4.6% from physicians – a nearly tenfold advantage. GPT-3.5’s responses were significantly longer than physicians’, averaging 211 words versus 52. A sensitivity analysis indicated that although longer physician messages garnered higher empathy ratings, they still fell short of the chatbot’s performance. Still, the authors caution that the extra length of chatbot responses might have been erroneously linked to higher perceived empathy. The sole study that favoured human HCPs was Reynolds (2024), which compared dermatologist vs. GPT-3.5 replies to patient-submitted dermatology queries; physician reviewers rated physicians as significantly more empathic (mean ~4 vs. ~3 for GPT-3.5; p = 0.001), whilst the difference noted by nonphysician reviewers did not reach statistical significance (p = 0.09). The SMD pooled across both reviewer groups was -0.99 (95% CI: -1.52 – -0.46). Overall, GPT-3.5 demonstrated higher empathy scores in three of the four studies, with high heterogeneity and a dermatology-specific exception.

*Subgroup: GPT-4 Narrative Synthesis*

Five studies (Guo, Yonatan-Leus, Soroudi, Armbruster, He) employed a 1–5 Likert scale. In Guo (2024), GPT-4’s responses to frequently asked thyroid-related patient questions from a hospital app were rated statistically significantly higher in compassion than those of both a junior and a senior specialist. For patient-reviewed scores, GPT-4 achieved a mean of 4.56 (SD = 0.35) compared to 3.82 (SD = 0.41) for junior specialists and 3.81 (SD = 0.47) for senior specialists (p < 0.001). For surgeon-reviewed scores, GPT-4 had a mean of 3.98 (SD = 0.62) versus 3.33 (SD = 0.52) for junior specialists and 3.33 (SD = 0.40) for senior specialists (p = 0.007). The SMD was 1.42 (95% CI: 0.85–1.99), encompassing both patient and surgeon reviewers and incorporating pooled values for GPT-4 and the combined junior / senior surgeon group. In Yonatan-Leus (2024), GPT-4’s “empathic concern” for mental health–related inquiries from social media exceeded that of mental-health practitioners with an effect size of 0.60, noted as large effect by the authors, and a SMD of 0.97 (CI: 0.73 – 1.21). Soroudi (2024) tested responses to common breast reconstruction questions from the electronic health record (EHR). Specific to GPT-4 alone, the model outperformed physicians and APPs (when pooled) by a SMD of 0.83 (95% CI: -0.09 – 1.75), with GPT-4’s “full” format achieved a raw mean empathy rating of 3.4 ± 0.5, exceeding the providers’ rating of 2 ± 0.9, whilst GPT-4’s “brief” (where it was prompted to ‘briefly explain’) format (2.1 ± 0.5) was closer to that of physicians/APPs. Armbruster (2024) likewise found GPT-4 to be significantly more empathic than physicians, whether judged by physician or patient reviewers, with the chatbot scoring 4.18 on a 1–5 scale compared to 2.70 for a web-based physician panel. Pooled across both reviewer types, the SMD was 1.44 (CI: 1.13 – 1.76). Notably, in this study, women patients rated GPT-4’s empathy slightly higher than men (4.46 vs 4.14; p=0.049), although both groups viewed the chatbot as more empathic than the human panel. He et al. (2024) also found GPT-4 to be significantly more empathic in responses to autism-related questions, reporting a mean empathy score of 3.64 (95% CI: 3.57–3.71) vs. 3.13 (95% CI: 3.04–3.21) for physicians, with an SMD of 0.80 (CI: 0.62 – 0.99). Furthermore, 62.34% of GPT-4’s replies scored 4 or higher, compared to 43.51% of physician responses.

Two subsequent studies (Xu and Yong) employed a 1–10 Likert scale. Xu (2024) investigated frequently asked questions about systemic lupus erythematosus (SLE) submitted to a patient education platform, finding that GPT-4’s answers were rated statistically significantly more empathic than rheumatologists’ overall, with a mean difference 0.46 (95% CI: 0.23–0.69) when measured by rheumatologists. The difference emerged clearly in Chinese, whereas English ratings showed no significant difference. From the patient perspective, GPT-4’s empathy was on par with rheumatologists in Chinese but significantly higher in English. The SMD was 0.23 (95% CI: -0.06 – 0.51), calculated across all combined ratings, pooling data from both language groups and rater panels. Yong (2024) had GPT-4 and patient relations department (PRD) staff to respond to patient complaints in a tertiary health institution and determined that GPT-4 was significantly more empathic, scoring 8 (IQR 7–9) vs. 4 (IQR 3–6), with an SMD of 2.08 (95% CI: 1.27 – 2.88). In another approach, Small (2024) compared drafts created by GPT-4 and HCPs from various backgrounds (physicians, nurses, and frontline staff) for outpatient internal medicine queries: primary care physicians (PCPs) first determined which drafts were “usable,” and amongst these, empathy was then assessed. GPT-4’s usable responses were rated as empathic in 37.2% of cases, compared with 16.5% for HCPs (a 126% relative increase), corresponding to a SMD of 0.48 (95% CI: 0.16 – 0.80). Linguistic analysis also revealed GPT-4’s more positive tone (mean positive polarity: 0.21 ± 0.14 vs. 0.13 ± 0.25; p = 0.02). Finally, Meyer et al. (2024) employed a 6-level rating scale to evaluate empathy in responses to laboratory-interpretation queries, observing GPT-4 to be “Excellent” in 22% and “Good” in 57% of cases, whereas physicians only reached 2% and 23%, respectively – a statistically significant difference in GPT-4’s favour and corresponding to a SMD of 1.44 (95% CI: 1.12 – 1.75).

Together, these nine studies consistently indicate that GPT-4 achieves higher empathy scores than human comparators across a broad range of clinical topics, rating scales, and evaluator backgrounds. Nonetheless, heterogeneity remains high – some contexts may yield smaller gains.

*Narrative Synthesis of Non-Meta-Analysed Results*

Chen et al. (2024) simultaneously assessed GPT-3.5 and GPT-4 and Claude on the same dataset for oncology queries, preventing distinct subgroup allocation (without cherry-picking or double counting). All three chatbots achieved higher empathy scores than oncologists for cancer-related inquiries from social media, with GPT-3.5 scoring 3.17 (95% CI: 3.10–3.24) and GPT-4 scoring 3.28 (95% CI: 3.20–3.36), compared to 2.43 (95% CI: 2.32–2.53) for oncologists. GPT-4 ranked highest among the chatbot models, followed by Claude and GPT-3.5. Li (2024) was also not included in meta-analysis, as it provided no specific metrics. They compared empathy-related language between human clinicians and Med-PaLM 2 using qualitative coding of conversation transcripts. Defining "empathy" through codes for appreciation, acknowledgment, and compassion, the study measured the average message count per conversation for each category. The analysis revealed that human clinicians used empathy-related language statistically significantly more frequently than the AI, although Med-PaLM 2 exhibited a slight edge in "compassion" markers.

The following three studies were included in the meta-analysis, however only the GPT-3.5/4 arms were included. We excluded the following models to avoid double-counting. Meyer (2024) also evaluated Gemini Pro and Le Chat (Mistral Large) against physicians; both AI models achieved a statistically significantly higher rating. Gemini was rated excellent in 31% and good in 40% of cases, Le Chat in 8% and 41%, and physicians only 2% and 23%, respectively. He (2024) also assessed ERNIE Bot, which slightly underperformed compared to physicians, scoring 3.11 (95% CI, 3.04–3.18) vs. 3.13 (95% CI, 3.04–3.21) on a 1-5 Likert Scale respectively, although this was not statistically significant. Soroudi (2024) also evaluated GPT-3; its “full” format achieved a mean empathy rating of 3.7 ± 0.6, exceeding the providers’ rating of 2 ± 0.9, while GPT-3’s “brief” format (2.4 ± 0.6) was closer to that of physicians/APPs.

Overall, the collective findings indicate that multiple AI models frequently achieve higher empathy ratings than human clinicians, although variability exists among models and specific measures of empathic communication.
