## Supplementary material for "AI Chatbots Versus Human Healthcare Professionals: A Systematic Review and Meta-Analysis of Empathy in Patient Care": Multimedia Appendix D Risk of Bias Assessment

### Multimedia Appendix D. Risk of Bias Assessment for 15 Included Studies

Given that the majority of the included studies are non-randomised (e.g., cross-sectional, comparative analyses), the ROBINS-I tool was selected. This tool is specifically designed to assess the risk of bias in non-randomised studies of interventions: <https://methods.cochrane.org/robins-i>

**Maida et al. (2024)**

**Article Title:** ChatGPT vs. neurologists: a cross‑sectional study investigating preference, satisfaction ratings and perceived empathy in responses among people living with multiple sclerosis.

**Citation:** Maida E. et al. (2024). *Journal of Neurology*, 271:4057–4066. <https://doi.org/10.1007/s00415-024-12328-x>

| **Domain** | **Assessment** | **Judgment** |
| --- | --- | --- |
| **Confounding** | The study adjusted for several covariates including age, sex, treatment duration, clinical descriptors, cognitive deficits, depressive symptoms, and educational attainment. However, the manner in which the four questions were chosen (i.e., curated from patients’ emails, then narrowed by neurologists) may introduce confounding. The selected FAQs may not represent a random sample of all possible patient queries, and the wording (or domain) of those questions might differentially favour ChatGPT vs. a neurologist’s style. | **Moderate** |
| **Selection of Participants** | Empathy reviewers (patient-proxies) were recruited through digital communication platforms with a response rate of 39.7%. This may lead to selection bias as responders may differ from non-responders (e.g., people who are more digitally engaged might be more likely to respond). There is no comparison of responders vs. non-responders. | **Moderate** |
| **Classification of Interventions** | AI (ChatGPT) and human (neurologist) responses were clearly distinguished and appropriately categorised. However, the paper notes that ChatGPT responses had a slightly more informal tone, so some participants might suspect which was AI, potentially introducing bias (but this is an inherent difference between AI and humans and *arguably* can’t be controlled). | **Low** |
| **Deviation from Intended Interventions** | No deviations from intended interventions were reported; both AI and neurologist responses were generated and presented consistently. | **Low** |
| **Missing Data** | The study does not indicate any missing data issues. | **Low** |
| **Measurement of Outcomes** | They used the CARE scale for empathy, which is validated, albeit slightly modified for a digital context. Its use in a purely text-based “Q&A” scenario is less typical but still plausibly relevant. | **Low** |
| **Selection of the Reported Result** | The study focused on main outcomes (preference, satisfaction, empathy) without evidence of selective reporting of results. | **Low** |

**Xu et al. (2024)**

**Article Title:** ChatGPT4’s proficiency in addressing patients’ questions on systemic lupus erythematosus: a blinded comparative study with specialists.

**Citation:** Xu D. et al. (2024). *Rheumatology*, 63:2450–2456.

| **Domain** | **Assessment** | **Judgment** |
| --- | --- | --- |
| **Confounding** | The questions were curated (from “kidney online platform”) rather than selected at random. There may be a degree of selection bias in the two researchers choosing questions, but this is reduced through two researchers. No mention of covariates; age, sex, severity of SLE, etc.; they were not accounted for. Because the questions were public, it makes it difficult to obtain these patient characteristics. However, there is no evidence to suggest serious confounding. | **Moderate** |
| **Selection of Participants** | A subset of five rheumatologists answered these questions. They appear to be conveniently sampled (from specific hospitals in China, Hong Kong, and the USA). Variation in their typical style, thoroughness, etc. could influence outcome scores. | **Moderate** |
| **Classification of Interventions** | Responses, both AI-generated and derived from rheumatologists, were randomized and anonymised to ensure unbiased evaluation | Low |
| **Deviation from Intended Interventions** | No deviations from intended interventions were reported; both AI and rheumatologist responses were generated and presented consistently. | Low |
| **Missing Data** | The paper does not indicate that any question–answer pairs were omitted post hoc; they appear to have included all 95 curated questions with a complete set of ChatGPT4 and rheumatologist answers. There is no indication of participant (rater) drop-out or missing data. To account for potential non-response or incomplete data, authors increased the target sample size by 10% | Low |
| **Measurement of Outcomes** | The empathy instrument was not a fully validated measure—rather a 0–10 rating for multiple domains, including empathy. This reliance on a bespoke scoring method could introduce subjectivity and differences in how the raters interpret “empathy.” | Moderate |
| **Selection of the Reported Result** | Focussed on 6 main outcomes (scientific validity, logical consistency, comprehensibility, completeness, satisfaction level and empathy), with no evidence of selective reporting. Results are shown by subgroup: Chinese vs. English. | Low |

**Guo et al. (2024)**

**Article Title:** Comparing ChatGPT’s and Surgeon’s Responses to Thyroid-related Questions From Patients.

**Citation:** Guo S. et al. (2024). *The Journal of Clinical Endocrinology & Metabolism*, 00:1–10. <https://doi.org/10.1210/clinem/dgae235>

| **Domain** | **Assessment** | **Judgment** |
| --- | --- | --- |
| **Confounding** | No mention of confounders or covariates (e.g. patient age) in the paper. Only stratification was for seniority of surgeon (i.e. senior or junior specialist), and for different hospital locations. The researches ‘worked with patients to organise the top 40 questions’, these were further screened by one surgeon against exclusion criteria, resulting in 28 curated questions. This selection bias could introduce bias if the selected questions systematically favour one type of response over another. Nonetheless these were rigorous methods in question selection to ensure that the questions are relevant and representative of common patient inquiries, with no evidence for serious confounding. | Moderate |
| **Selection of Participants** | The study used a convenience sample of 26 patients (all from one hospital) and 11 surgeons (some from the same hospital, some from outside). They do not clarify in detail how patients were recruited – only that they had to be “able to read/write and willing to participate”. No discussion of how many were invited vs. how many declined. If only a certain subset of patients (e.g., those more comfortable using technology or more enthusiastic about research) volunteered, that could bias the perception of ChatGPT’s empathy. Similarly, surgeons volunteering might have systematically different attitudes to AI. | Moderate |
| **Classification of Interventions** | Responses were blinded, however the vastly different response lengths (ChatGPT produced many more words – as noted by the authors) might still cue some raters. Yet the authors did remove certain ChatGPT “boilerplate” statements. Overall, the classification of who generated which response seems to have been adequately blinded. | Low |
| **Deviation from Intended Interventions** | No deviations from intended interventions were reported; both AI and surgeons’ responses were generated and presented consistently. | Low |
| **Missing Data** | They included 37 valid questionnaires (11 surgeons + 26 patients). They do not report high rates of incomplete questionnaires, only that they excluded forms if ≥10 answers were identically scored. There is no sign of systematic dropout or large amounts of missing data. | Low |
| **Measurement of Outcomes** | Reported on three main outcomes: compassion, accuracy, and comprehensiveness. Used a 5-point Likert scale: not validated. | Moderate |
| **Selection of the Reported Result** | No evidence of selective reporting: reported on outcomes as stated. | Low |

**Yonatan-Leus & Brukner (2024)**

**Article Title:** Comparing perceived empathy and intervention strategies of an AI chatbot and human psychotherapists in online mental health support.

**Citation:** Yonatan-Leus, R., & Brukner, H. (2024). *Counselling and Psychotherapy Research*, 00, 1–9. <https://doi.org/10.1002/capr.12832>

| **Domain** | **Assessment** | **Judgment** |
| --- | --- | --- |
| **Confounding** | No explicit mention of confounders (e.g., age of patients). The data is public and anonymous and thus it is difficult/impossible to obtain these patient characteristics. Reddit is an informal platform, and the nature of patient-clinician interactions here likely does not reflect the formality of real clinical contexts. Licensed professionals on Reddit (unpaid) may not always behave as they would in a formal therapy setting. The responses were gathered this way. They are offering ad-hoc, often short-form replies. These volunteers on the platform possibly systematically differ from a more general group of mental-health professionals. It is highly plausible that licensed therapists do not “perform” empathy as thoroughly as they would in a real or more formal setting. | Serious |
| **Selection of Participants** | 150 questions from the ‘Ask A Therapist’ subreddit. May be a degree of responder bias, as those that post questions are more likely to be engaged with their mental health. The selection process could be non-random or convenience-based. They excluded certain question types (e.g., “ethical queries” or “personal experience requests”), which may systematically shape the final set. | Moderate |
| **Classification of Interventions** | AI and human psychotherapists’ responses were adequately separated. Researchers and participants blinded. | Low |
| **Deviation from Intended Interventions** | No deviations from intended interventions were reported; both AI and psychotherapists’ responses were generated and presented consistently | Low |
| **Missing Data** | Based on the study details provided, it appears that the dataset is complete—with 300 responses analysed and no indication of gaps in the data | Low |
| **Measurement of Outcomes** | Investigated perceived empathic concern and perspective-taking. This was based on a scale from 1 to 5, with definitions and examples of each one provided. This wasn’t validated. | Moderate |
| **Selection of the Reported Result** | No evidence of selective reporting | Low |

**Ayers et al. (2023)**

**Article Title:** Comparing physician and artificial intelligence chatbot responses to patient questions posted to a public social media forum.

**Citation:** Ayers JW. et al. (2023). *JAMA Internal Medicine*, 183(6):589–596. <https://doi.org/10.1001/jamainternmed.2023.1838>

| **Domain** | **Assessment** | **Judgment** |
| --- | --- | --- |
| **Confounding** | The study uses patient questions and physician responses from Reddit's r/AskDocs forum, which may not accurately represent typical patient-physician interactions in clinical settings. Authors acknowledged that “actual physicians may form answers based on established patient-physician relationships”. Volunteer Physicians: Responders on Reddit are unpaid volunteers, (arguably) impacting motivation, effort levels, and communication tone from those working in a professional clinical setting. However, they don’t acknowledge or address these issues, which could have a large impact on quality of responses, and thus it may not be a fair comparison. Authors could have obtained a sample of some replies from clinical settings and compared them to see if the data for Reddit is a fair representation. | Serious |
| **Selection of Participants** | 195 questions from r/AskDocs. The authors had previously noted (<https://www.youtube.com/watch?v=ECzx49KmPtA&ab_channel=ExternalMedicinePodcast>) that many using this platform cannot afford healthcare and thus turn to Reddit for free advice, consequently this data is likely from those from a poorer economic background. This is selection bias – those who can afford traditional healthcare are likely excluded. | Serious |
| **Classification of Interventions** | Physician and AI responses were adequately separated and classified. Blinding was performed. | Low |
| **Deviation from Intended Interventions** | No deviations from intended interventions were reported; both AI and physicians’ responses were generated and presented consistently | Low |
| **Missing Data** | No evidence of significant missing data or incomplete question–answer pairs. | Low |
| **Measurement of Outcomes** | Primary outcomes were “the quality of information provided” and “the empathy or bedside manner provided”. Measured using a Likert scale; not validated. | Moderate |
| **Selection of the Reported Result** | No evidence of selective reporting | Low |

**D. et al. (2024)**

**Article Title:** Comparing Provider and ChatGPT Responses to Breast Reconstruction Patient Questions in the Electronic Health Record.

**Citation:** D. et al. (2024). *Annals of Plastic Surgery*, 93(5):541–545. <https://doi.org/10.1097/SAP.0000000000004090>

| **Domain** | **Assessment** | **Judgment** |
| --- | --- | --- |
| **Confounding** | No explicit mention of confounders. The set of questions was intentionally selected (i.e., curated) from a large pool. This selection process could confound performance. | Moderate |
| **Selection of Participants** | Ten questions were ultimately chosen after screening 1623 queries for frequency, context, and complexity. Although that process is described, it’s not random; thus, it’s prone to selection bias (the final 10 might over-represent certain complexities or omit mundane questions). | Moderate |
| **Classification of Interventions** | Both AI and plastic surgeon responses were adequately classified. Blinding performed to minimise bias. | Low |
| **Deviation from Intended Interventions** | No deviations from intended interventions were reported; both AI and surgeons’ responses were generated and presented consistently | Low |
| **Missing Data** | They appear to have collected ratings for all question–answer pairs, with no mention of dropouts or missing evaluations. Sample size is small but apparently complete. | Low |
| **Measurement of Outcomes** | Used 1-5 Likert scale to measure empathy and accuracy. Non-validated.  Readability was assessed using the Flesch Reading Ease, which is validated. | Moderate |
| **Selection of the Reported Result** | No evidence of selective reporting of results | Low |

**Reynolds et al. (2024)**

**Article Title:** Comparing the quality of ChatGPT- and physician-generated responses to patients’ dermatology questions in the electronic medical record.

**Citation:** Reynolds K. et al. (2024). *Clin Exp Dermatol*, 49:715–718. <https://doi.org/10.1093/ced/llad456>

| **Domain** | **Assessment** | **Judgment** |
| --- | --- | --- |
| **Confounding** | No explicit mention of confounders. The authors do mention “randomly extracting” 40 patient questions from the EMR (electronic medical record), eventually including 31 in the final analysis. However, there was still no true randomisation of the ‘intervention’. The messages extracted from EMR were not randomly assigned to one intervention or another (AI or human). Here, the physician replies were generated in an actual clinical scenario, while ChatGPT replies were generated after the fact. There is no random assignment of which “arm” is delivering the real-world reply. | Moderate. |
| **Selection of Participants** | Many using this platform cannot afford healthcare and thus turn to Reddit for free advice | Low |
| **Classification of Interventions** | Reponses adequately classified as either AI or physician responses. Blinding performed to minimise bias. | Low |
| **Deviation from Intended Interventions** | No deviations from intended interventions were reported; both AI and physicians’ responses were generated and presented consistently | Low |
| **Missing Data** | They started with 40 question–answer pairs and ended up with 31. This was driven by certain exclusions: e.g., questions that lacked meaningful content. While some might question whether that could bias the analysis, the final set was fully evaluated by the 10 raters, and the study does not mention further missing data once a question was included. | Low |
| **Measurement of Outcomes** | Answers rated on ‘overall quality’, ‘readability’, ‘accuracy’, ‘thoroughness’ and ‘level of empathy’ based on 1-5 Likert scale. Non-validated | Moderate |
| **Selection of the Reported Result** | No evidence of selective reporting of results | Low |

**Meyer et al. (2024)**

**Article Title:** Comparison of ChatGPT, Gemini, and Le Chat with physician interpretations of medical laboratory questions from an online health forum.

**Citation:** Meyer, et al. (2024). *Clinical Chemistry and Laboratory Medicine*.

| **Domain** | **Assessment** | **Judgment** |
| --- | --- | --- |
| **Confounding** | Addresses some such as age and gender (presumably gleaned from the userr’s own statement in the post), and word count. Residual confounders likely remain. The study compares physician responses on Reddit’s r/AskDocs with AI chatbot responses. Physicians on Reddit do not have an established patient–physician relationship, and usually have only the textual detail the user provided (similar to the chatbots). Nonetheless, confounding arises if the volunteer physicians have different motivations, time constraints, or styles compared to standard (more formal) clinical settings (which is likely). | Serious |
| **Selection of Participants** | All data come from r/AskDocs, which is inherently self-selected. This is not necessarily representative of patients in everyday medical practice. Many using this platform cannot afford healthcare and thus turn to Reddit for free advice (as discussed: <https://www.youtube.com/watch?v=ECzx49KmPtA&ab_channel=ExternalMedicinePodcast>) as so are of a poorer economic background. The paper reports user ages (median 27) and gender, presumably gleaned from the user’s own statement in the post. This might or might not accurately reflect actual patient demographics and is likely much younger due the digital nature of the platform. | Serious |
| **Classification of Interventions** | Responses were adequately classified as either AI generated (different chatbots) or physician generated. Blinding performed to minimise identification bias. | Low |
| **Deviation from Intended Interventions** | No deviations from intended interventions were reported; both AI and physicians’ responses were generated and presented consistently | Low |
| **Missing Data** | The authors ended with 100 posts from an initial 635, applying certain exclusion criteria. This is fairly large. We do not see mention of partial dropouts after the main selection, or missing outcome data for the “quality” metrics. So while the selection might not be representative, there is no indication that missing data within the included set systematically biased results. | Low |
| **Measurement of Outcomes** | The authors did anonymise and remove any AI disclaimers, so presumably the raters were blinded to which was the chatbot vs. the physician (though style differences might still inadvertently unblind them). The outcome measures (quality, clarity, empathy, and “ranking”) are subjective. They used “1–6” or “1–4” ordinal scales. The two-physician panel tried to resolve disagreements via consensus. This reduces but does not eliminate subjectivity. Because many of these constructs (like empathy or clarity) lack a standardized, validated scale, some subjectivity remains | Moderate |
| **Selection of the Reported Result** | No evidence of selective reporting of results | Low |

**Li, Wang, Strachan, et al. (2024)**

**Article Title:** Conversational AI in health: Design considerations from a Wizard-of-Oz dermatology case study.

**Citation:** Li, Wang, Strachan, et al. (2024). *Extended Abstracts of the CHI Conference on Human Factors in Computing Systems (CHI EA ’24)*. https://doi.org/10.1145/3613905.3651891

| **Domain** | **Assessment** | **Judgment** |
| --- | --- | --- |
| **Confounding** | No explicit mention of confounders. Some potential confounders mentioned (age, ethnicity), but no mention of how these were dealt with. The authors state participants were assigned randomly to either the Clinician Agent (n=10) or the Supervised LLM Agent (n=8). This is helpful for reducing some confounding. However, there is no mention of stratification or attempts to balance disease severity or complexity of skin conditions, which might affect how participants perceive the chatbot’s responses. A potential confounder is that the LLM conversation was intentionally supervised by a dermatologist to ensure “safe and accurate” responses; that workflow itself might differ in intensity between participants and might systematically shape the outcome. In addition, the sponsor’s involvement may or may not influence study design in ways unaccounted for. | Moderate |
| **Selection of Participants** | Participants were recruited in the USA through a third-party organization that had existing relationships with individuals eligible for the study. Eligibility criteria for the study included having an existing skin concern and the desire to learn more information about it, regardless of whether or not the skin concern had previously been examined by a healthcare professional.  No mention of randomisation, leading to selection bias. No mention of how ‘third party organisation’ recruits individuals. Small sample size (n=18), leading to decreased power of study. | Moderate |
| **Classification of Interventions** | This was a Wizard-of-Oz design: participants did not know the random condition. The authors say participants were under the impression they were interacting with an automated chatbot in both arms. In the “Clinician Agent,” a dermatologist was actually typing all responses. In the “Supervised LLM” condition, the LLM produced an output that was then *vetted* (and possibly edited) by a dermatologist. The labelling is straightforward. However, the role of dermatologist supervising LLM agent introduces bias, as the LLM isn’t completely autonomous, impacting a direct comparison. | Serious |
| **Deviation from Intended Interventions** | The design is quite controlled: participants typed questions into a chat, and behind the scenes a dermatologist or an LLM+dermatologist responded. Because it is a Wizard-of-Oz format, the authors had full control over the “exposure.” There is no mention of participants or clinicians deviating from the assigned approach. | Low |
| **Missing Data** | The authors do not report major dropout or incomplete rating data. They mention 18 participants completed the immediate post-interaction surveys, and presumably, each provided chat logs. A 2-week follow-up survey was conducted; presumably, not everyone might have responded, but the paper states results in aggregate without noting major attrition that would cause large bias. | Low |
| **Measurement of Outcomes** | The researches counted how many messages featured empathy-related codes, e.g. “I’m sorry to hear that”. They employed ad hoc codes (“appreciation,” “acknowledgment,” “compassion”) rather than using a standardised clinical empathy scale (e.g., CARE measure, Jefferson Scale of Empathy, etc.). This can introduce subjectivity into which phrases get categorised as empathy. Certain phrases might appear empathic in isolation but lack genuine empathic intent within the larger conversation; Frequency ≠ Depth or Quality of Empathy. Additionally, the authors are from Google evaluating a Google-based model. That raises at least the potential for sponsor bias or confirmation bias in how qualitative data are interpreted (though we do not see strong evidence they concealed negative data). | Moderate |
| **Selection of the Reported Result** | No evidence of selective reporting of results. Although it could be argued that there may be a greater risk of this with qualitative analysis, the authors minimised this risk through inductive approach, rather than deductive. They share chat counts, empathy discussion, concerns about “lack of empathy” in the LLM agent. So they do not appear to be downplaying negative aspects. | Low |

**Armbruster et al. (2024)**

**Article Title:** “Doctor ChatGPT, Can You Help Me?” The Patient’s Perspective: Cross-Sectional Study.

**Citation:** Armbruster J. et al. (2024). *J Med Internet Res*, 26:e58831. <https://doi.org/10.2196/58831>

| **Domain** | **Assessment** | **Judgment** |
| --- | --- | --- |
| **Confounding** | Identified some potential covariates, including sex, speciality, age that may affect outcomes. Likely other residual confounders remain. The study might have confounding if certain question complexities or specialties systematically favor one responder. The authors tried to mitigate that by random sampling (20 questions per specialty) and including multiple specialties. | Moderate |
| **Selection of Participants** | The authors used convenience sampling (“convenient sampling took place from in-hospital patients and patients entering the outpatient department of a tertiary care hospital”). Some also participated from partner practices. This may be subject to selection bias. | Moderate |
| **Classification of Interventions** | Responses were adequately classified as either AI or physician. Blinding performed to minimise identification bias | Low |
| **Deviation from Intended Interventions** | The two sets of answers (ChatGPT vs. EP) were compiled into packages. The participants rated them in a cross-sectional, one-time session. There is no sign that participants or the investigators deviated from the stated design. | Low |
| **Missing Data** | They report that 64 patient raters completed 200 question packages total (though the text says “200 packages were completed by patients” from 64 unique participants). Each package included 10 question–answer pairs, so presumably they got 2000 total ratings for each arm. That suggests minimal missingness overall. The authors do not report major attrition or incomplete data. | Low |
| **Measurement of Outcomes** | Patients were asked about empathic rating/friendliness, and if the response to the question would have helped. Physicians were asked same questions, plus if the information was professionally correct, and if it contains potentially harmful advice. Measured on 1-5 Likert scale; not validated. | Moderate |
| **Selection of the Reported Result** | No evidence of selective reporting of results | Low |

**Small et al. (2024)**

**Article Title:** Large Language Model–Based Responses to Patients’ In-Basket Messages.

**Citation:** Small et al. (2024). *JAMA Network Open*.

| **Domain** | **Assessment** | **Judgment** |
| --- | --- | --- |
| **Confounding** | No explicit mention of confounders; however, subgroup analyses performed on whether response quality varied with HCP type (physicians and nonphysicians) and  patient message classification (laboratory results, medication refill requests, paperwork, and general medical advice). The survey design used “branching logic”—respondents first decided if the message was “usable,” then were asked about empathy, personalization, etc. That means empathy was only assessed *if* the rater already found the response “usable”. This can confound empathy judgments with “usefulness”. The effect is that empathy is not systematically measured for all responses, so poor (or “unusable”) responses never even get an empathy rating, which might inflate perceived empathy for certain arms or produce skewed estimates. | Moderate |
| **Selection of Participants** | A convenience sample of 16 PCPs (from a large internal medicine listserv of 1189) responded. This 1.3% response rate is low, raising questions about selection bias: those more curious or enthusiastic about AI might respond. The study uses real EHR messages from pilot clinics, but many messages were excluded if they required external context. That might skew the sample toward simpler or more self-contained queries. | Moderate |
| **Classification of Interventions** | Responses were adequately classified as either AI or healthcare professional responses. Blinding performed to minimise identification bias | Low |
| **Deviation from Intended Interventions** | No deviations from intended interventions were reported; both AI and HCP’s responses were generated and presented consistently | Low |
| **Missing Data** | They do not mention systematic dropout after rating certain messages. Some message sets were never shown if external context was needed, but that is more about selection than missing data. | Low |
| **Measurement of Outcomes** | Empathy is measured only for “usable” responses—so empathy is not assessed at all for nonusable messages. This can systematically overrepresent empathy among whichever arm’s messages end up being considered “usable”. That is arguably a design flaw or potential measurement artifact, because many “poor” messages never get an empathy rating. The main scales (information quality, communication style, usability) are Likert-based and not validated. | Moderate |
| **Selection of the Reported Result** | No evidence of selective reporting of results | Low |

**Wan et al. (2024)**

**Article Title:** Outpatient reception via collaboration between nurses and a large language model: a randomized controlled trial.

**Citation:** Wan et al. (2024). *Nature Medicine*, 30:2878–2885. <https://doi.org/10.1038/s41591-024-03148-7>

| **Domain** | **Assessment** | **Judgment** |
| --- | --- | --- |
| **Confounding** | The authors mention they did simple 1:1 random allocation (by sealed envelopes) to either “nurse–SSPEC collaboration” or “nurse only”. The randomised controlled trial design minimizes confounding by evenly distributing known and unknown confounders between the nurse–SSPEC and nurse-only groups. Random allocation of participants to intervention arms addresses potential baseline differences. | Low |
| **Selection of Participants** | The authors excluded certain groups (patients with psychological disorders or who refused to sign consent). This might yield a more “cooperative” sample with potentially different baseline empathy perceptions. The final included sample (n=2,164) is fairly large, which strengthens generalizability, though it’s from a single centre and single reception site.  Post-randomisation dropout is low (21 total, 12 in nurse–SSPEC, 9 in nurse). It is not stated whether these dropouts might systematically differ in how they experience empathy. | Low |
| **Classification of Interventions** | Participants were randomised to two cohorts: the nurse–SSPEC (site-specific prompt engineering chatbot) collaboration group and the nurse group. However, the collaboration between nurses and SSPEC introduces complexity. The oversight by nurses may lead to higher empathy scores not solely attributable to the AI but to the combined human-AI interaction. This supervision can inadvertently enhance the perceived empathy of AI responses compared to purely human responses. This is an issue when assessing the intrinsic capabilities of the SSPEC without contextual influence of human oversight (rather than a flaw in how the RCT was conducted). | Serious |
| **Deviation from Intended Interventions** | The main potential risk is partial adoption of “AI style” by nurses who also worked with the SSPEC, or “learning contamination”, but the study does not mention seeing or controlling for that. The authors say 6 uncertain cases out of 1,080 had “false-negative alerts”, so the nurse–SSPEC model might have had “off-protocol” content in those few interactions. This does not appear to systematically bias empathy, just a small fraction. | Low |
| **Missing Data** | The paper says 2,185 were randomised, 21 dropped out, leaving 2,164. No mention that empathy data were missing. The final sample includes 1,080 vs 1,084 participants. The small dropout is balanced across arms, so no reason to suspect differential missingness for empathy. | Low |
| **Measurement of Outcomes** | The main empathy outcome is a Likert scale from 1 to 5. The authors do not use a validated empathy scale (like CARE or Jefferson). Instead, they used a custom dimension. This subjectivity is a potential bias. | Moderate |
| **Selection of the Reported Result** | No evidence of selective reporting of results. | Low |

**Yong et al. (2024)**

**Article Title:** Performance of Large Language Models in Patient Complaint Resolution: Web-Based Cross-Sectional Survey.

**Citation:** Yong L.P.X. et al. (2024). *J Med Internet Res*, 26:e56413. https://doi.org/10.2196/56413

| **Domain** | **Assessment** | **Judgment** |
| --- | --- | --- |
| **Confounding** | The authors attempt to measure some confounders (like word count, respondent being health care vs. non–health care worker). The authors do show that ChatGPT’s advantage persists even when controlling for word count in some analyses, but not all confounding variables are addressed. | Moderate |
| **Selection of Participants** | The authors mention that participants were “randomly selected and invited”, but they do not elaborate on how that random selection was done or whether certain groups were overrepresented. | Moderate |
| **Classification of Interventions** | Responses were adequately classified as either AI or human. Blinding performed to minimise identification bias | Low |
| **Deviation from Intended Interventions** | No major evidence that the study deviated from how they said they would present scenarios. | Low |
| **Missing Data** | The final sample was 188 participants who fully answered the questionnaires. The authors do not mention major dropouts or incomplete data. So, there is no sign of large missing outcome data. | Low |
| **Measurement of Outcomes** | The authors used a 10-point Likert scale with no midpoint, it was not a validated tool. | Moderate |
| **Selection of the Reported Result** | No sign of withheld outcomes or selective reporting. | Low |

**Chen et al. (2024)**

**Article Title:** Physician and Artificial Intelligence Chatbot Responses to Cancer Questions From Social Media.

**Citation:** Chen D. et al. (2024). *JAMA Oncology*, 10(7):956–960. https://doi.org/10.1001/jamaoncol.2024.0836

| **Domain** | **Assessment** | **Judgment** |
| --- | --- | --- |
| **Confounding** | Source of Physician Responses: The physician answers come from Reddit’s r/AskDocs forum, where volunteer responders do not have a preexisting patient–physician relationship, and where patients may have unique reasons for seeking free, online advice (e.g., difficulty affording health care). Mismatch With Real-World Clinical Practice: Physicians responding on Reddit are not in a formal clinical setting, possibly leading to different effort levels, tone, or thoroughness than in a standard patient–physician encounter. | Serious |
| **Selection of Participants** | This selection of 195 (or 200) question–answer pairs from an online forum is likely nonrepresentative. Patients who post on r/AskDocs differ demographically and socioeconomically from the general patient population, thus limiting generalisability. The authors do not attempt to compare or adjust for potential selection differences, so the sample may systematically bias outcomes for the AI vs. physician comparison. | Serious |
| **Classification of Interventions** | The authors do not explicitly detail the method used to blind or randomize the response identity to evaluators, although they do mention that each response was “blinded and randomly ordered”. | Low |
| **Deviation from Intended Interventions** | No deviations from intended interventions apparent. | Low |
| **Missing Data** | No evidence of large missing data or differential dropout. | Low |
| **Measurement of Outcomes** | They used a 5-point Likert scale – this isn’t’ validated. | Moderate |
| **Selection of the Reported Result** | No evidence of selective reporting of results | Low |

**He et al. (2024)**

**Article Title:** Physician Versus Large Language Model Chatbot Responses to Web-Based Questions From Autistic Patients in Chinese: Cross-Sectional Comparative Analysis.

**Citation:** He W. et al. (2024). *J Med Internet Res*, 26:e54706. https://doi.org/10.2196/54706

| **Domain** | **Assessment** | **Judgment** |
| --- | --- | --- |
| **Confounding** | The study involves autistic patients seeking medical advice on DXY (a web-based medical consultation platform). Autism severity, patient background, and region might systematically influence the types of questions asked. No mention of controlling for patient-level factors such as age, socioeconomic status, or severity of ASD symptoms. | Moderate |
| **Selection of Participants** | 100 patient consultation samples from a from DXY—a widely acknowledged, web-based, medical consultation platform. Those using DXY may differ from the general population with ASD (e.g., more digital literacy, more resources). | Moderate |
| **Classification of Interventions** | Responses were adequately classified as either AI or physician. Blinding performed to minimise identification bias | Low |
| **Deviation from Intended Interventions** | No deviations from intended interventions were reported. The authors systematically collected the physician responses from DXY and generated chatbot replies by inputting the original queries, so each question consistently had 1 physician answer, 1 ChatGPT answer, and 1 ERNIE Bot answer. | Low |
| **Missing Data** | They selected 239 questions in total, with 717 total evaluations (each question having 3 responses: from a physician, ChatGPT, and ERNIE Bot). The study does not mention major dropout or partial data omission for those 239 questions, so it appears all question–answer sets were fully evaluated. The authors do not explicitly discuss missing data handling, but from the final numbers, it appears minimal (all 717 evaluations are reported). | Low |
| **Measurement of Outcomes** | 4 dimensions: relevance, accuracy, usefulness, and empathy. They used a Likert scale to rate each one 1-5 (not validated). | Moderate |
| **Selection of the Reported Result** | No evidence of selective reporting of results | Low |

**Total Risks:**

1. **Maida et al. (2024):** Moderate
2. **Xu et al. (2024):** Moderate
3. **Guo et al. (2024):** Moderate
4. **Yonatan-Leus & Brukner (2024):** Serious
5. **Ayers et al. (2023):** Serious
6. **D. et al. (2024):** Moderate
7. **Reynolds et al. (2024):** Moderate
8. **Meyer et al. (2024):** Serious
9. **Li, Wang, Strachan, et al. (2024):** Serious
10. **Armbruster et al. (2024):** Moderate
11. **Small et al. (2024):** Moderate
12. **Wan et al. (2024):** Serious
13. **Yong et al. (2024):** Moderate
14. **Chen et al. (2024):** Serious
15. **He et al. (2024):** Moderate
