## Supplementary material for "AI Chatbots Versus Human Healthcare Professionals: A Systematic Review and Meta-Analysis of Empathy in Patient Care": Multimedia Appendix E Study-Specific Results

**Armbruster (2024)**

- **Patient-Reviewed Scores:**
  - ChatGPT-4: 4.18 vs. Expert Panel (EP): 2.7
- **Physician-Reviewed Scores:**
  - ChatGPT-4: 4.49 vs. Expert Panel (EP): 3.07

**Ayers (2023)**

- **Empathy Scores:**
  - ChatGPT-3.5: 3.65 (95% CI, 3.55–3.75)
  - Physicians: 2.15 (95% CI, 2.03–2.27)
- **Evaluator Preference:**
  - Chatbot responses were preferred in 78.6% (95% CI: 75.0–81.8%) of evaluations.
- **Proportion of Empathic Responses:**
  - Chatbot: 45.1% (95% CI: 38.5–51.8%)
  - Physicians: 4.6% (95% CI: 2.1–7.7%)
    *(Note: Chatbot responses were ~9.8 times more likely to be rated empathetic.)*

**Chen (2024)**

- **Overall Empathy Score (Cognitive + Emotional):**
  - Physicians: 2.43 (95% CI, 2.32–2.53)
  - Chatbot 1 (GPT-3.5): 3.17 (95% CI, 3.10–3.24)
  - Chatbot 2 (GPT-4): 3.28 (95% CI, 3.20–3.36)
  - Chatbot 3 (Claude): 3.62 (95% CI, 3.53–3.70)

**Guo (2024)**

- **Patient-Reviewed:**
  - *Junior Surgeons:* AI – 4.56 ± 0.35 vs. Human – 3.82 ± 0.41
  - *Senior Surgeons:* AI – 4.56 ± 0.35 vs. Human – 3.81 ± 0.47
- **Surgeon-Reviewed:**
  - *Junior Surgeons:* AI – 3.98 ± 0.62 vs. Human – 3.33 ± 0.52
  - *Senior Surgeons:* AI – 3.98 ± 0.62 vs. Human – 3.33 ± 0.40

**He (2024)**

- **Empathy Scores (Likert 1–5):**
  - ChatGPT-4: 3.64 (95% CI, 3.57–3.71)
  - ERNIE Bot: 3.11 (95% CI, 3.04–3.18)
  - Physicians: 3.13 (95% CI, 3.04–3.21)

**Li (2024)**

- **Observations:**
  - Human clinicians used more frequent “appreciation” and “acknowledgment” markers overall.
  - AI responses included slightly more “compassion” statements (e.g., “I’m sorry to hear that”).
    *(Specific numeric metrics were not provided.)*

**Maida (2024)**

- **Sub-question Scores (as reported):**
  - Q1: ChatGPT – 34.24 vs. Neurologist – 28.91
  - Q2: ChatGPT – 31.25 vs. Neurologist – 30.89
  - Q3: ChatGPT – 32.38 vs. Neurologist – 31.58
  - Q4: ChatGPT – 29.54 vs. Neurologist – 30.87
    *(Note: A single overall sub-scale sum was not provided.)*

**Meyer (2024)**

- **ChatGPT:**
  - Excellent: 22%
  - Good: 57%
  - Satisfactory: 20%
  - Sufficient: 1%
  - Poor: 0%
- **Gemini Pro:**
  - Excellent: 31%
  - Good: 40%
  - Satisfactory: 21%
  - Sufficient: 6%
  - Poor: 0%
  - Inadequate: 2%
- **Le Chat:**
  - Excellent: 8%
  - Good: 41%
  - Satisfactory: 48%
  - Sufficient: 3%
  - Poor: 0%
- **Physicians:**
  - Excellent: 2%
  - Good: 23%
  - Satisfactory: 38%
  - Sufficient: 32%
  - Poor: 5%
    *(Data retrieved from study appendices.)*

**Reynolds (2024)**

- **For Physician Reviewers:**
  - Physicians: 4 (SD < 1) vs. ChatGPT-3: 3 (SD < 1)
- **For Non-Physician Reviewers:**
  - Physicians: 4 (SD < 1) vs. ChatGPT-3: 3 (SD < 1)

**Small (2024)**

- **Empathic Response Counts:**
  - ChatGPT: 32 out of 86 messages (37.2%)
  - HCPs: 13 out of 79 messages (16.5%)
  - *(Relative increase: 125.5%)*
- **Linguistic Drivers (mean ± SD):**
  - *Subjectivity:* ChatGPT – 0.54 (0.16) vs. HCP – 0.31 (0.23) *(p < 0.001)*
  - *Positive Polarity:* ChatGPT – 0.21 (0.14) vs. HCP – 0.13 (0.25) *(p = 0.02)*
    *(No additional empathy statistics were reported.)*

**Soroudi (2024)**

- **Provider Scores:**
  - Physicians: 2.1 ± 1.0
  - APPs: 2.0 ± 0.8
  - Combined Providers: 2.0 ± 0.9
- **ChatGPT Versions:**
  - ChatGPT-3 Full: 3.7 ± 0.6
  - ChatGPT-4 Full: 3.4 ± 0.5
  - ChatGPT-3 Brief: 2.4 ± 0.6
  - ChatGPT-4 Brief: 2.1 ± 0.5
  - Combined Chatbot (all versions): 2.9 ± 0.8
  - Combined Brief Chatbot: 2.3 ± 0.5

**Wan (2024)**

- **Internal Validation (n = 7,084):**
  - “SPPEC”: 4.12 ± 0.86 vs. Nurse: 3.39 ± 1.21
- **Randomized Controlled Trial:**
  - “SSPEC” (supervised): 4.14 ± 0.98 vs. Nurse-only: 3.27 ± 1.22

**Xu (2024)**

- **Chinese Evaluations:**
  - *Rheumatologist:* AI – 7.84 ± 0.69 vs. Human – 7.36 ± 0.59
  - *SLE Patients:* AI – 9.56 ± 0.67 vs. Human – 9.38 ± 0.95
- **English Evaluations:**
  - *Rheumatologist:* AI – 4.68 ± 2.37 vs. Human – 4.55 ± 2.04 *(p = 0.801)*
  - *SLE Patients:* AI – 8.55 ± 0.98 vs. Human – 7.23 ± 1.21

**Yonatan-Leus (2024)**

- **Results:**
  - ChatGPT-4: 4.17 ± 0.70
  - Human (Mental Health Licensed Professionals): 3.40 ± 0.87

**Yong (2024)**

- **Results (using a 1–10 scale):**
  - ChatGPT-4: Median 8 (IQR 7–9)
  - Human (Patient Relations Officers): Median 4 (IQR 3–6)
